# Rare coding and noncoding variants map 1,342 diseases and biomarkers in 490,549 whole genomes

**DOI:** 10.64898/2026.03.24.26349148

**Authors:** Yuxin Yuan, Yuanyuan Guan, Yannuo Feng, Tony Chen, Yingzi Zhang, Baoqun Chang, Shijie Fan, Chang Lu, Wenyuan Li, Xiaoyu Li, Xihao Li, Xihong Lin, Zilin Li

## Abstract

Rare genetic variants are increasingly recognized as important contributors to human trait architecture, with noncoding variants accounting for a substantial portion of the heritability. These variants tend to be less polygenic and more biologically specific than common variants, remaining understudied across large biobanks. Here we analyzed whole genome sequencing (WGS) data from up to 490,549 UK Biobank participants to assess the effects of rare coding and noncoding variants across 1,342 phenotypes, including 944 diseases, 76 clinical biomarkers, and 322 metabolomics traits. We developed and applied *STAARpipelinePheWAS*, a scalable framework for WGS phenome-wide rare variant association analysis, identifying 49,121 genome-wide significant gene-trait pairs. Our study presents a comprehensive map of noncoding rare variant associations in both disease and biomarker domains. Many associations were undetected in prior exome- or array-based studies and were enriched in drug targets and biologically coherent pathways. All results are publicly accessible through an interactive portal (https://www.staarphewas.org/), offering a foundational resource for rare variant discovery, functional interpretation, and translational genomics.

## Introduction

The discovery of disease-associated variants has accelerated the development of effective and safe therapeutics^1–3^. Drug targets supported by human genetic evidence are substantially more likely to progress through clinical development, with a 2.6-fold higher probability of regulatory approval compared with targets lacking such evidence^4–6^. For example, the identification of rare loss of function variants in *PCSK9* associated with low-density lipoprotein cholesterol levels led to clinical trials and the subsequent approval of *PCSK9* inhibitors for treating cardiovascular disease^7,8^.

Genome-wide association studies (GWAS) have identified tens of thousands of common and low-frequency variants associated with human diseases and biomarker traits^9^. These findings have enabled systematic mapping of pleiotropy and genetic correlations, supporting data-driven disease stratification, and revealing shared biological mechanisms that link biomarkers and disease risk^9,10^. However, GWAS also has several limitations. First, association signals often localize near highly pleiotropic genes, making it difficult to pinpoint trait-specific biological mechanisms^11^. Second, common and low-frequency variants explain only a fraction of pedigree-based narrow-sense heritability, leaving a substantial proportion unaccounted for^12,13^. Rare variants (RVs) could help address these gaps. Collectively, they account for a sizable share of pedigree-based heritability^12^ and exhibit larger biological effects than common variants^14,15^. Rare variants often implicate genes and pathways with direct causal roles in disease, providing clearer mechanistic insight and more precise opportunities for therapeutic development^16–18^. Importantly, RVs are more likely to highlight trait-specific genes rather than highly pleiotropic loci^11^, offering a complementary and biologically focused perspective on human disease genetics.

Whole-exome sequencing (WES) studies have enabled systematic discovery of rare coding variant associations, but it surveys only a small fraction of ∼2-3% of the genome that encodes protein-coding exons and excludes nearly all noncoding variation^19–21^.

Whole-genome sequencing (WGS) studies overcome this limitation by extending RV discovery into noncoding regulatory regions. WGS-based heritability analyses in the UK Biobank indicate that WGS data can capture the majority of pedigree-based heritability across complex traits that recovers much of the previously “missing” heritability, with roughly 20% of the total attributable to rare variants, and that coding and noncoding variants account for approximately 20% and 80% of rare variant heritability, respectively^12^. By incorporating noncoding regulatory elements, WGS provides access to mechanisms of gene regulation that cannot be evaluated through WES or array-based studies^21^. Although rare noncoding variation is increasingly recognized as an important contributor to human diseases and complex traits, it remains relatively underexplored at the biobank scale^20–24^. In addition, WGS identifies more than 10% of functional coding variation than exome sequencing based technologies^18,19^, further enhancing its utility for disease-associated rare coding variant detection. With the emergence of large-scale, deeply phenotyped WGS resources such as the UK Biobank, it is feasible to systematically characterize the contributions of both rare coding and noncoding variants across a wide range of diseases and clinically relevant biomarkers^19^.

Here we leveraged WGS data from up to 490,549 UK Biobank participants, developed and applied the *STAARpipelinePheWAS* framework to conduct a phenome-wide association study (PheWAS) of rare coding and noncoding variants across 1,342 phenotypes, including 944 diseases, 76 clinically biomarkers, and 322 metabolomic traits. We identified 682 coding and 239 noncoding gene-trait pairs for diseases, and 16,195 coding and 32,005 noncoding pairs for biomarkers and metabolomic traits, respectively, capturing both well-established and previously unreported associations. These results provide a comprehensive resource of coding and noncoding genetic associations (https://www.staarphewas.org/), and illustrate how WGS-based rare variant analysis can uncover mechanisms underlying diseases and biomarkers with direct relevance for target discovery and translational genomics.

## Results

### Study Design

We analyzed WGS data from up to 490,549 UK Biobank participants. Participants had a median age of 58 years (interquartile range 50-63), and 54.24% were female. We conducted a large-scale PheWAS to evaluate rare coding and noncoding associations across 944 diseases defined using Phecodes^25^ and 398 quantitative biomarkers (**Fig. 1a**, **Supplementary Tables 1-2** and **Methods**). Disease phenotypes were grouped into 16 categories, with circulatory system diseases representing the largest category, followed by genitourinary (**Supplementary Figure 1a, Supplementary Table 1** and **Methods**). The median number of cases per category ranged from 976 to 6,272, with digestive diseases contributing to the largest case counts (**Supplementary Figure 1b**). Biomarkers included 76 clinical measurements spanning blood, urine, and physical traits, together with 322 nuclear magnetic resonance (NMR) metabolomic traits (**Supplementary Table 2** and **Methods**). These biomarkers were classified into 11 categories, with NMR lipoprotein subclasses forming the largest group (**Supplementary Figure 1c-d**).

**Fig. 1.**
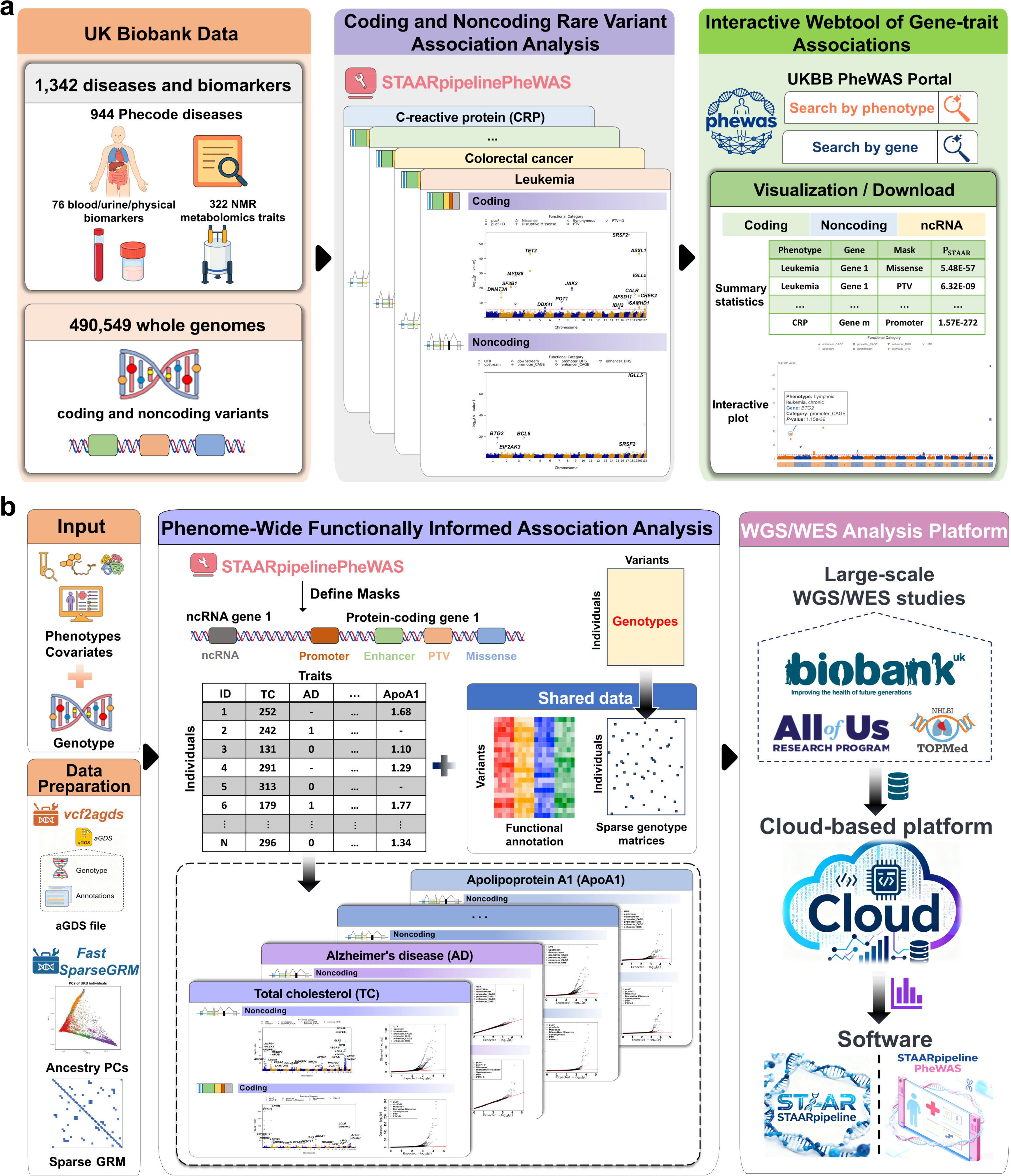
Study design schematic and overview of the cloud-based whole-genome sequencing (WGS) analysis workflow. **(a)** Study design schematic. This figure summarizes the analytical framework used to assess the contribution of rare coding and noncoding variants to 1,342 diseases and biomarkers in up to 490,549 UK Biobank genomes. Variant set analyses for rare variants (MAF < 1%) were performed using *STAARpipelinePheWAS*, including gene-centric analysis of protein-coding genes using 7 coding variant masks and 7 noncoding variant masks, and one mask of ncRNA genes. An accompanying interactive web portal (https://www.staarphewas.org/) provides a resource for exploring rare variant contributions to human traits and diseases. **(b)** Overview of the cloud-based WGS analysis workflow. (i) Prepare input data, including genotypes, phenotypes, and covariates. (ii) Convert WGS data from VCF to annotated Genomic Data Structure (aGDS) format using *vcf2agds* toolkit, and compute ancestry PCs and a sparse GRM using *FastSparseGRM*; (iii) Perform phenome-wide, functionally informed rare variant association analyses using *STAARpipelinePheWAS*, which achieves computational efficiency through sparse genotype matrix operations and single-pass extraction of genotypes and annotations shared across multiple phenotypes. (iv) The workflow supports cloud execution for WGS and WES analyses at biobank scale.

To analyze this wide range of phenotypes in 490,549 UK Biobank genomes, we developed *STAARpipelinePheWAS* (STAARpipeline for Phenome-Wide Association Analysis), a scalable framework for phenome-wide functionally informed rare variant association analysis in WGS and WES studies (**Fig. 1b**, **Supplementary Note** and **Methods**). For coding RV analysis, *STAARpipelinePheWAS* provides seven coding functional categories (masks) of protein-coding genes. For noncoding RV analysis, *STAARpipelinePheWAS* provides seven noncoding masks for regulatory regions of protein-coding genes, and one mask for noncoding RNA (ncRNA) genes (**Methods**). The framework supports simultaneous analysis of multiple phenotypes and achieves computation efficiency by directly extracting and manipulating sparse genotype matrices throughout the workflow and extracting genotypes and functional annotations in a single pass across phenotypes (**Fig. 1b**).

For coding RV analysis, we applied Bonferroni-corrected genome-wide significance thresholds of *α* = 0.05/(20,000 × 7) = 3.57 × 10^−7^ accounting for 7 different masks across ∼20,000 protein-coding genes. For noncoding RV analysis of protein-coding genes, we applied Bonferroni-corrected genome-wide significance thresholds of *α* = 0.05/(20,000 × 7) = 3.57 × 10^−7^ accounting for 7 different noncoding masks. For ncRNA genes, we used a Bonferroni-corrected genome-wide significance thresholds of *α* = 0.05/20,000 = 2.50 × 10^−6^ accounting for ∼20,000 ncRNA genes^26,27^.

### Rare coding genetic variants associated with diseases

We identified 1,687 genome-wide significant coding associations for 944 diseases (**Fig. 2a**, **Supplementary Table 3**), corresponding to 682 unique gene-trait pairs. Among these associations, the mask of protein-truncating and disruptive missense RVs accounted for the largest proportion of 23.18%, whereas synonymous RVs contributed the smallest fraction of 3.08% (**Fig. 2b**). The detected gene-trait pairs were particularly abundant in neoplasms, endocrine/metabolic, hematologic, and circulatory system diseases (**Supplementary Table 4**). Of the 682 gene-trait pairs, 310 (45.45%) overlapped with associations reported in previous GWAS studies^28^, and 362 (53.08%) were identified in previous large-scale exome-wide analyses of UK Biobank WES or WGS data^15,19^. Notably, 171 gene-trait pairs (25.07%) were not detected in either prior array- or sequencing-based studies, underscoring the added discovery value of our coding RV analysis (**Supplementary Table 4**).

**Fig. 2.**
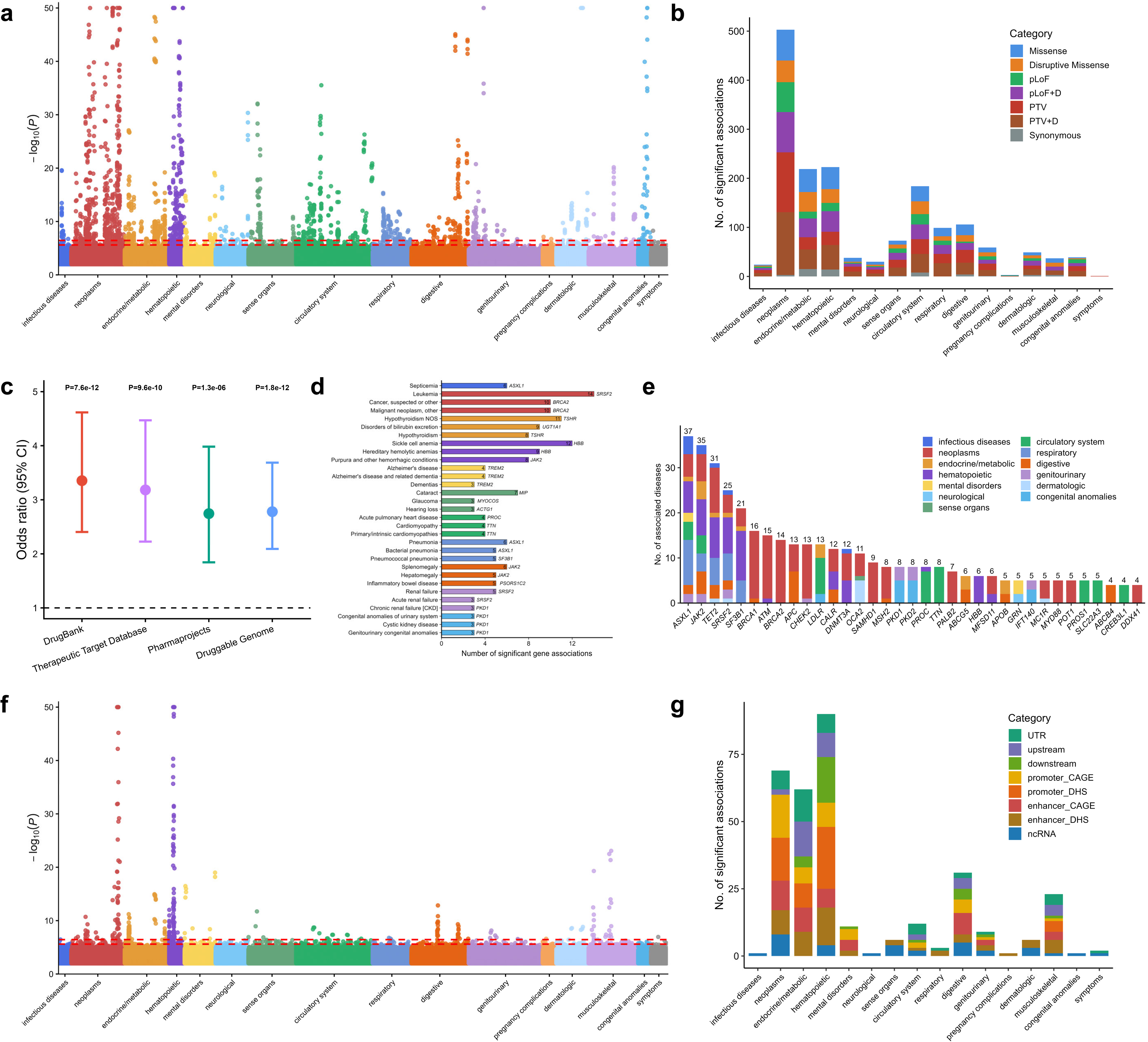
Summary of rare coding and noncoding genetic variants associated with 944 diseases. **a** Gene-centric coding analyses (7 masks) for diseases, colored by disease category. Each column on the x-axis represents a trait, and each point represents a mask-trait association. The dashed lines indicate two genome-wide significance thresholds (*α* = 0.05/20,000 = 2.50 × 10^−6^ and *α* = 0.05/(20,000 × 7) = 3.57 × 10^−7^). The y-axis is capped at −log_10_(*P*) = 50, and associations with *P* < 0.01 are shown. **b** Frequency of mask-trait associations across the 7 coding masks, grouped by disease category. **c** Enrichment of FDA-approved drug targets among protein-coding genes identified in coding analyses. Error bars represent 95% confidence intervals (CIs). Four drug-target databases were used: DrugBank, Therapeutic Target Database (TTD), Informa Pharmaprojects, and the Druggable Genome. *P* values were calculated using two-sided Fisher’s exact tests. **d** Top three diseases with the largest numbers of significant gene-trait pairs from coding analyses within each disease category, colored by category. Gene labels correspond to the most significant association (minimum *P* value) for each disease. Diseases with at least three significant gene associations are shown. **e** Stacked bar chart summarizing disease roles of protein-coding genes identified in coding analyses, grouped by disease category. Bars represent the top 35 genes with the greatest number of significant gene-trait associations, with numbers above each bar indicating the total number of associated diseases. **f** Gene-centric noncoding analyses (8 masks) for diseases, colored by disease category. Each column on the x-axis represents a trait, and each point represents a mask-trait association. The dashed lines indicate two genome-wide significance thresholds (*α* = 0.05/20,000 = 2.50 × 10^−6^ and *α* = 0.05/(20,000 × 7) = 3.57 × 10^−7^). The y-axis is capped at −log_10_(*P*) = 50, and only associations with *P* < 0.01 are shown. **g** Frequency of mask-trait associations across 8 noncoding masks grouped by disease category.

The identified coding RV associations were significantly enriched for known drug targets (DrugBank odds ratio (OR): 3.35 (95% CI: 2.40-4.62), *P* = 7.58 × 10^−12^; Therapeutic Target Database (TTD) OR: 3.18 (95% CI: 2.23-4.47), *P* = 9.62 × 10^−10^) (**Fig. 2c**, **Methods**). Canonical disease-associated genes were robustly recovered, with examples including *TSHR* with hypothyroidism, *HBB* with hereditary hemolytic anemias, *TREM2* with Alzheimer’s disease, and *TTN* with cardiomyopathy^15,29–31^, supporting the validity of our analytical framework (**Fig. 2d**). *ASXL1* exhibited the largest number of associations, with 37 traits across diverse disease categories (**Fig. 2e**, **Supplementary Table 5**), including neoplasms, hematopoietic disorders, and respiratory diseases, consistent with its broad involvement in epigenetic regulation across multiple biological systems. Other genes with coding RVs associated with multiple diseases include *JAK2*, *TET2*, *BRCA1*, *BRCA2*, *ATM*, *APC*, and *CHEK2*, many of which are implicated in cancer or hereditary disease syndromes.

In addition to known disease-associated genes, previously unreported association signals were also identified. For example, disruptive missense RVs in *SF3B1* showed strong associations with hematologic malignancies, including myeloproliferative disease (*P* = 3.12 × 10^−50^) and leukemia (*P* = 9.34 × 10^−22^), consistent with its known somatic role in myeloid neoplasms^32,33^. Beyond these known associations, broader associations were also observed with platelet and leukocyte disorders, including aplastic anemia (*P* = 2.19 × 10^−7^), thrombocytopenia (*P* = 3.31 × 10^−10^), and neutropenia (*P* = 1.76 × 10^−9^), all of which are replicated at the nominal significance level of 0.05 in the All of Us dataset^34^. These patterns suggest that disruption of *SF3B1* function influences multiple hematologic traits, consistent with its role in blood cell and platelet homeostasis. Additionally, protein-truncating variants in *PSORS1C1* and *PSORS1C2*, which are genes within the major histocompatibility complex (MHC) linked psoriasis susceptibility 1 (*PSORS1*) locus, were associated with inflammatory bowel disease (*P* = 7.63 × 10^−9^ for *PSORS1C1*; *P* = 9.86 × 10^−10^ for *PSORS1C2*) and ulcerative colitis (*P* = 1.67 × 10^−8^ for *PSORS1C1*; *P* = 3.83 × 10^−9^ for *PSORS1C2*), consistent with shared genetic architecture between autoimmune skin and intestinal conditions^35–37^.

### Rare noncoding variants reveal regulatory mechanisms of diseases

In parallel, we identified 328 genome-wide significant noncoding associations for 944 diseases (**Fig. 2f**, **Supplementary Table 6**), and 58.84% are detected by enhancer or promoter masks (**Fig. 2g**). These associations represent 239 unique gene-trait pairs, which were most prevalent in neoplasms and hematologic traits (**Supplementary Table 7**). Among the 239 noncoding gene-trait pairs, 131 pairs (54.81%) overlapped with associations reported in previous GWAS studies^28^, 18 pairs (7.53%) were identified in prior sequencing-based studies^15,19^, and 43 pairs (17.99%) overlapped with associations in our coding RV analysis (**Supplementary Table 7**). Notably, 95 of the 239 gene-trait pairs (39.75%) were not detected in any of these three categories, underscoring the distinct contribution of noncoding regulatory variants to the genetic architecture of diseases.

These noncoding associations both expanded known biology and revealed under-recognized contributors to diseases (**Supplementary Table 8**). For example, upstream RVs of *TNF* were associated with celiac disease (*P* = 1.42 × 10^−13^). Although *TNF* is a well-established immune regulator and therapeutic target for inflammatory diseases^38^, its genetic association with celiac disease has not been reported. Promoter DHS RVs in *BCL2* were associated with lymphocytic leukemia (*P* = 2.25 × 10^−9^) and immune-related disorders (*P* = 1.16 × 10^−7^). Despite its role as a central regulator of apoptosis and an approved oncology target in hematologic malignancies^38^, the association between *BCL2* and these phenotypes has not been observed in prior genetic association studies, including those based on common variant from GWAS and rare variant from sequencing studies^15,19^.

Beyond regulatory regions of protein coding genes, we also identified associations involving rare variants in noncoding RNA loci. ncRNA RVs in *BGLT3* were associated with sickle cell anemia (*P* = 1.34 × 10^−9^), a signal not reported in earlier genetic association studies. Functional evidence from multiple systems supports the relevance of this finding^39,40^. We also detected RVs in *MIR184* were associated with corneal opacity and related corneal disorders. While not identified in previous population-scale association studies, *MIR184* has been implicated in congenital anterior segment dysgenesis through family-based studies^41^ and in Fuchs endothelial corneal dystrophy in a recent case-only genetic study^42^.

### Rare coding and noncoding variant associations in cancer susceptibility

To illustrate how our results inform the genetic architecture of human diseases, we examined cancers as a representative disease group. Across 83 cancer traits, we identified 503 coding and 69 noncoding associations, corresponding to 203 and 49 unique gene-trait pairs, respectively (**Fig. 3a**). Among the 203 coding pairs, 159 (78.33%) had supporting evidence in previous GWAS^28^ or sequencing studies^15,19^ (**Supplementary Table 4**). In contrast, 24 of the 49 noncoding gene-trait pairs (48.98%) were not reported in previous GWAS and sequencing-based studies, nor were they observed in our coding RV analysis (**Supplementary Table 7**), highlighting the distinct contribution of noncoding regulatory variation. For example, promoter RVs in *BTG2* were associated with chronic lymphoid leukemia (*P* = 1.15 × 10^−36^), consistent with functional studies showing that *BTG2* regulates leukemic cell quiescence and chemoresistance^43^.

**Fig. 3.**
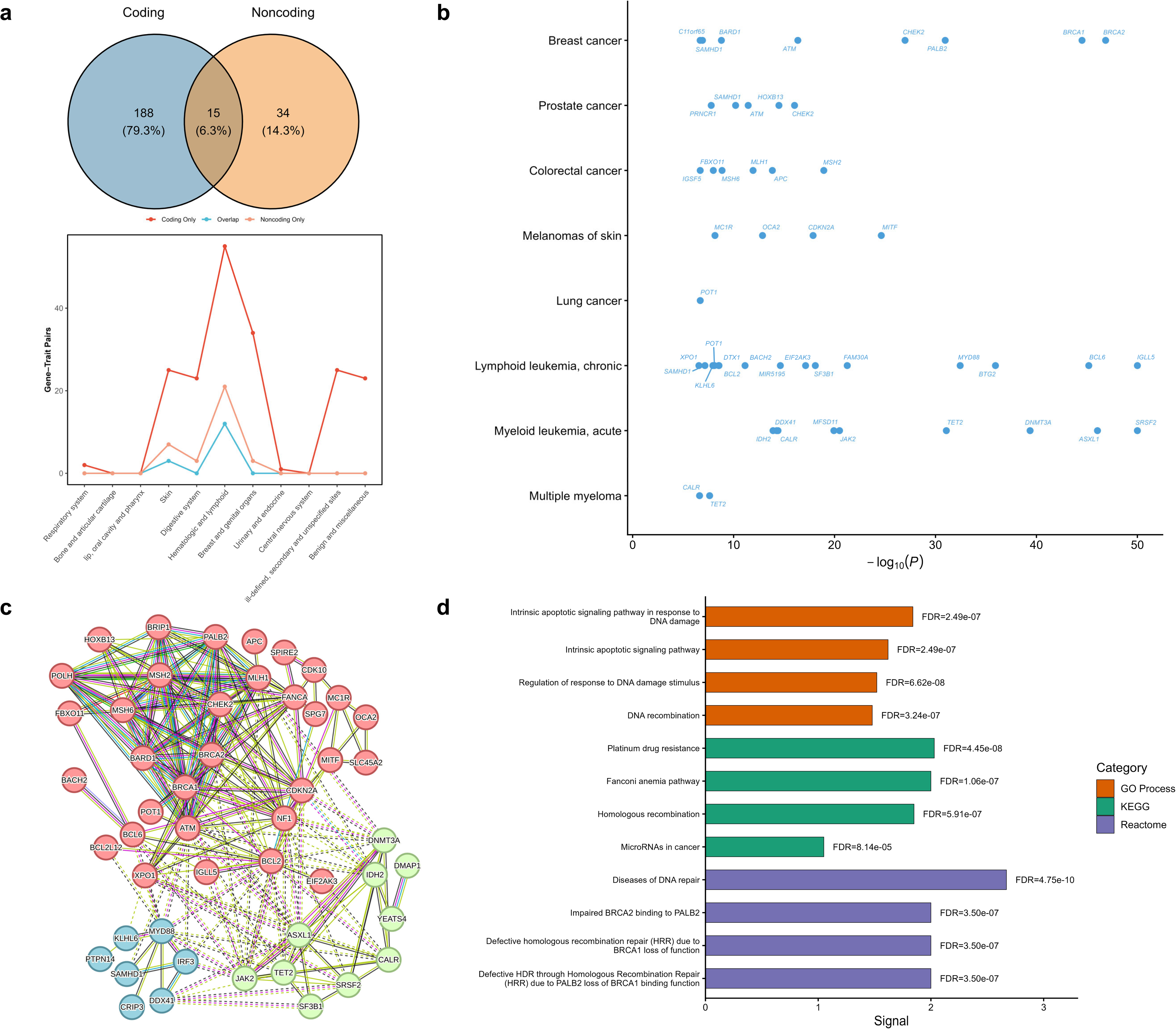
Summary of rare coding and noncoding genetic variants associated with 83 cancers. **a** Comparison of the number of gene-trait pairs identified in coding versus noncoding analyses. **b** Significant gene-trait pairs from coding and noncoding analyses across representative major tumor types. For each cancer, when a gene is associated through multiple masks, the association with the minimum *P* value is shown. The x-axis is capped at −log_10_(*P*) = 50. **c** Protein-protein interaction network of the 63 protein-coding genes identified across both coding and noncoding analyses. Orphan genes (i.e., genes without connections in the PPI network) were hidden. **d** Pathway enrichment analysis (GO, KEGG, Reactome) for the 63 identified protein-coding genes, showing the top four significantly enriched pathways from each database.

As expected, hematologic cancers showed dense clusters of associations involving established driver genes such as *JAK2*, *TET2*, *CALR*, *ASXL1*, *SRSF2* (**Fig. 3b**). Classic cancer susceptibility genes (*BRCA1*, *BRCA2*, *CHEK2*, *ATM*, *PALB2*) were associated with breast and ovarian cancers, while *CDKN2A*, *MC1R*, and *MITF* were linked to melanoma and other skin cancers. Fewer significant signals were detected for solid tumors, including associations involving *ATM* with stomach and pancreatic cancers and *POT1* with lung cancer. These findings are consistent with known cancer biology and demonstrate the robustness of our analytical framework.

The detected gene-trait pairs involved 63 protein-coding genes across 48 cancer traits. To investigate their functional relationships, we conducted a protein-protein interaction (PPI) analysis, which revealed an interconnected structure with three major gene clusters (**Fig. 3c** and **Methods**). Cluster 1 included genes involved in DNA repair and platinum drug resistance pathways (for example, *BRCA1, BRCA2*, *ATM*). Cluster 2 captured epigenetic and signaling regulation (for example, *JAK2*, *TET2*). Cluster 3 included genes implicated in immune and inflammatory responses (for example, *IRF3*, *MYD88*). These modules reflect key biological processes underlying cancer susceptibility. Functional enrichment analyses of the identified genes further supported their mechanistic relevance (**Fig. 3d**, **Supplementary Table 9** and **Methods**). We observed significant enrichment in pathways related to DNA damage response, DNA recombination, DNA repair, platinum drug resistance, Fanconi anemia pathway, homologous recombination, and impaired *BRCA2* binding to *PALB2*.

In addition to regulatory variation near protein coding genes, we also detected significant associations involving noncoding RNA genes. Rare variants in *FAM30A* and *MIR5195* were associated with chronic lymphoid leukemia. *FAM30A* has previously been implicated as a risk factor in acute myeloid leukemia, where higher expression correlates with poorer prognosis^44–46^. We also identified associations between RVs in *PRNCR1* (prostate cancer associated non-coding RNA 1) and prostate cancer, and between RVs in *RNU6-61P* and both liver cancer and intrahepatic bile duct cancer. These findings expand the scope of cancer-associated loci beyond protein coding genes and highlight the contribution of ncRNA genes to cancer susceptibility, components largely missed in coding gene-focused analyses.

### Rare coding variants reveal widespread biomarker associations

Serum and urine biomarkers are widely used to monitor health, diagnose disease, and guide treatment decisions^47–50^. To evaluate the contribution of rare coding variants contribution to biomarkers, we analyzed 398 biomarkers, including 76 clinical measures and 322 NMR metabolomic traits (**Supplementary Table 2** and **Methods**). In total, we identified 39,568 genome-wide significant coding associations (**Supplementary Fig 2a**, **Supplementary Table 10**). Missense RVs accounted for the largest proportion (27.54%), followed by the mask of protein truncating and disruptive missense RVs (18.97%), while synonymous RVs represented the smallest fraction (9.56%; **Supplementary Fig 2b**). These biomarker-associated genes were significantly enriched for known drug targets, suggesting therapeutic potential across diverse biomarker domains (**Fig. 4a**, **Supplementary Fig 3** and **Methods**).

**Fig. 4.**
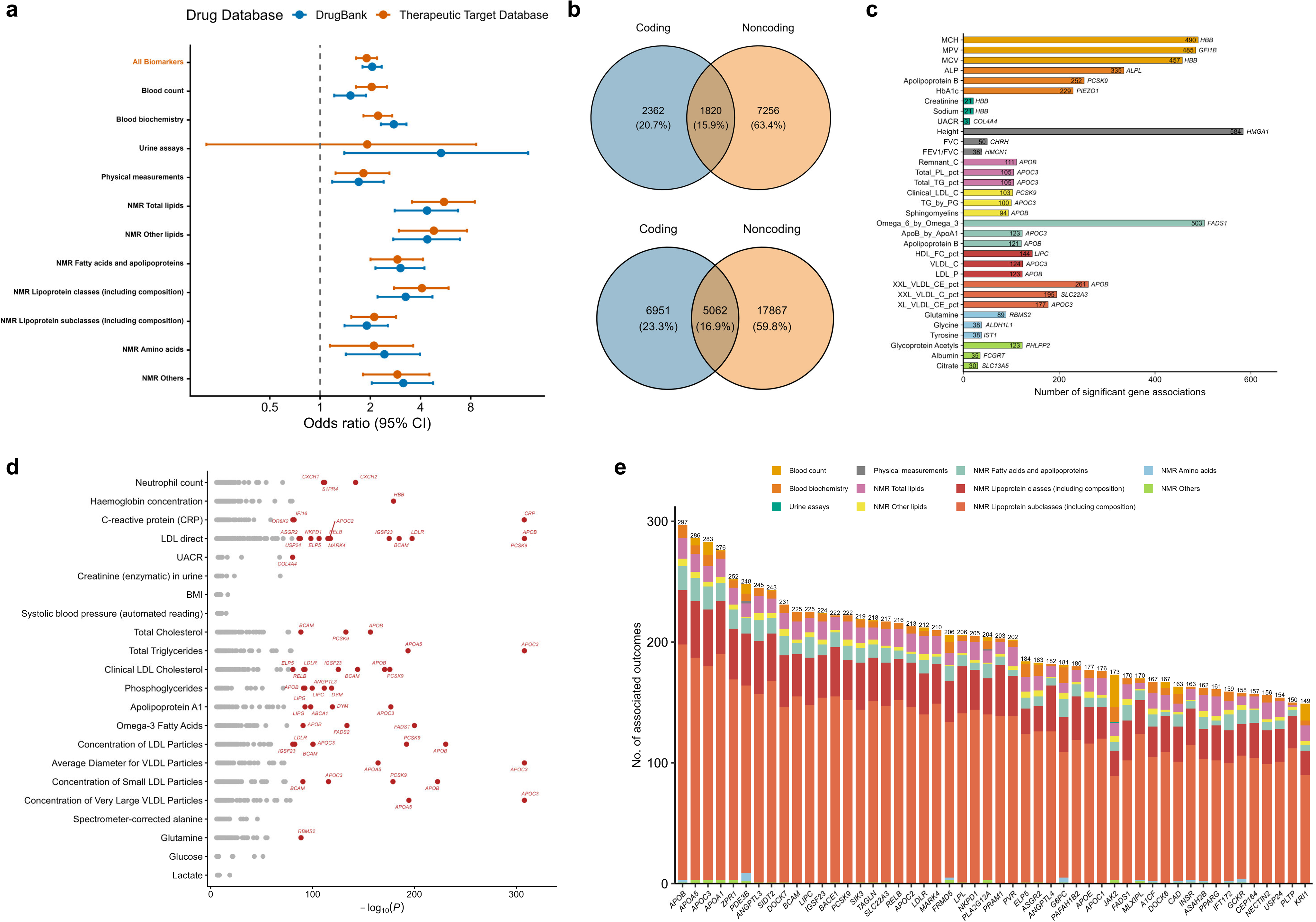
Summary of rare coding and noncoding genetic variants associated with 398 biomarkers. **a** Enrichment of FDA-approved drug targets among protein-coding genes identified from coding analyses. Error bars represent 95% CIs. DrugBank and Therapeutic Target Database (TTD) were used as drug-target reference databases. *P* values were calculated using two-sided Fisher’s exact tests. The first row represents enrichment across all 398 biomarkers. **b** Comparison of the number of gene-trait pairs identified in coding versus noncoding analyses for 76 clinical biomarkers (top) and 322 metabolomic traits (bottom). **c** Top three biomarkers within each biomarker category showing the largest numbers of significant gene-trait pairs identified across coding and noncoding analyses. Bars are colored by biomarker category, and gene labels correspond to the most significant association (minimum *P* value) for each biomarker. Biomarkers with at least three significant gene associations are included. Mean corpuscular haemoglobin (MCH), MCH; Mean platelet (thrombocyte) volume (MPV), MPV; Mean corpuscular volume (MCV), MCV; Alkaline phosphatase (ALP), ALP; Glycated haemoglobin (HbA1c), HbA1c; Creatinine (enzymatic) in urine, Creatinine; Sodium in urine, Sodium; urinary albumin-to-creatinine ratio, UACR; Forced vital capacity (FVC, best measure), FVC; FEV1/FVC ratio Z-score, FEV1/FVC. See Supplementary Table 2 for abbreviations of NMR metabolites. **d** Significant gene-trait pairs from coding and noncoding analyses for two representative biomarkers in each biomarker category. For biomarkers with multiple mask-specific associations, the minimum *P* value per gene is shown. Genes with *P* < 1 × 10^−80^ are labeled. **e** Stacked bar chart summarizing biomarker roles of protein-coding genes identified across coding and noncoding analyses, grouped by biomarker category. Shown are the top 50 genes with the highest numbers of significant gene-trait associations; numbers above bars denote the total number of associated biomarkers.

For the 76 clinical biomarkers, we identified 8,949 coding significant associations, corresponding to 4,182 unique gene-trait pairs (**Fig. 4b**). Among these, 3,781 pairs (90.41%) overlapped with associations reported in previous GWAS studies^28^, and 1,044 pairs (24.96%) were found in previous large-scale exome-wide analyses of UK Biobank WES or WGS data^15,19^. Notably, 315 gene-trait pairs (7.53%) were not detected in either prior array- or sequencing-based studies. For example, beyond its well-established role in blood cancers, disruptive missense RVs in *JAK2* were associated with polipoprotein A (*P* = 3.05 × 10^−35^), apolipoprotein B (*P* = 4.20 × 10^−39^), estimated glomerular filtration rate (eGFR) (*P* = 3.51 × 10^−22^), and urinary albumin-to-creatinine ratio (UACR) (*P* = 4.77 × 10^−9^), consistent with mouse model data implicating *JAK2* in lipid metabolism^51,52^ and kidney function^53^.

For the 322 NMR metabolomic traits, we identified 30,619 coding associations, corresponding to 12,013 unique gene-trait pairs (**Fig. 4b**). Among these, 7,017 pairs (58.41%) overlapped with previous GWAS findings^28^, and 4,127 pairs (34.25%) were observed in exome-wide analyses of UK Biobank WES or WGS data^15,19,48,50,51^.

Notably, 3,649 pairs (30.38%) were not detected in any of these prior studies. Illustrative examples include missense RVs in *A1CF*, associated with 158 NMR traits (81.01% not previously reported), and protein-truncating and disruptive missense RVs in *G6PC* associated with 166 NMR traits (60.24% not previously reported). These findings align with experimental studies linking these genes to apolipoprotein B, glycerol, and glucose metabolism^54,55^.

### Rare noncoding variants uncover regulatory contributions to biomarker variation

To complement coding analyses, we examined noncoding RVs across the same 398 biomarker traits. In total, we identified 56,025 significant noncoding associations (**Supplementary Fig 2c**, **Supplementary Table 11**), with the largest shares from enhancer masks (41.81%) and promoter masks (28.37%) (**Supplementary Fig 2d**).

For the 76 clinical biomarkers, we detected 15,762 noncoding associations, corresponding to 9,076 unique gene-trait pairs (**Fig. 4b**). Among these, 8,072 pairs (91.14%) overlapped with previous GWAS findings, 506 pairs (5.58%) overlapped with prior sequencing-based studies^15,19^, and 1,820 pairs (20.05%) overlapped with significant coding associations from our own analysis. Overall, 732 gene-trait pairs (8.07%) were not detected in any of these three categories. For example, enhancer RVs in *CALML6* were associated with 14 blood count traits, including red blood cell count previously reported by GWAS^28^, as well as previously unreported findings such as basophil, eosinophil, neutrophil, monocyte, and total white blood cell counts.

For the 322 NMR metabolomic traits, we identified 40,263 noncoding associations, corresponding to 22,929 gene-trait pairs (**Fig. 4b**). Among these, 14,655 pairs (63.91%) overlapped with associations previous GWAS findings, 2,234 pairs (9.74%) overlapped with prior sequencing-based studies^15,19,48,50,51^, and 5,062 pairs (22.08%) overlapped with significant coding associations in our own analysis. In total, 6,804 of the 22,929 gene-trait pairs (29.67%) were not detected in any of these three categories. Illustrative examples include enhancer RVs in *TYW1B*, associated with 115 NMR traits (85.22% not previously reported), and ncRNA RVs in *RNU4-80P* associated with 100 NMR traits (79.00% not previously reported).

### Integrated insights into biomarker biology and therapeutic relevance

As expected, we recapitulated well-established associations^11,15^, including *HBB* for blood count and urine assays, *NPR2* and *HHIP* for height, *PCSK9*, *APOB* and *APOC3* for lipid traits, and *CRP* for C-reactive protein, supporting the validity of our approach (**Fig. 4c-d**, **Supplementary Tables 12-14**, **Supplementary Fig 4**). Among all biomarkers, *APOB* showed the largest number of 297 associations, followed by other lipids-related genes such as *APOA5*, *APOC3*, *LIPC*, and *PCSK9* (**Fig. 4e**, **Supplementary Tables 12-14**, **Supplementary Fig 4**).

To better understand the functional relevance of biomarker-associated genes, we performed enrichment and PPI analyses using all identified protein-coding genes from both coding and noncoding analyses. These analyses further revealed coherent biological pathways, reinforcing the interpretability and functional relevance of the associations (**Supplementary Figs 5-15, Supplementary Tables 15-25**, **Supplementary Note**). These results highlight new opportunities for trait-specific mechanistic insights and therapeutic development across clinical and metabolomic domains.

## Discussion

We conducted a large-scale WGS-based PheWAS of rare coding and noncoding variants in 490,549 UK Biobank participants across 1,342 diseases and biomarkers. To enable analysis at this scale, we developed *STAARpipelinePheWAS*, a scalable and efficient framework that supports simultaneous functionally informed rare variant association analysis of multiple phenotypes. In total, we identified 682 coding and 239 noncoding gene-trait pairs for diseases, and 16,195 coding and 32,005 noncoding pairs for clinical biomarkers and metabolomic traits. These associations include both well-established and previously unreported findings and demonstrate the value of WGS in characterizing the rare variant architecture of human diseases and traits. Our results revealed significant enrichment of associated genes among known therapeutic targets, reinforcing the role of human genetics in drug discovery and validation. Protein-protein interaction networks and functional enrichment analyses highlighted biologically coherent pathways, supporting the interpretability and functional relevance of the associations. The identification of coding and regulatory signals across thousands of traits underscores the utility of integrating both types of variants in PheWAS to illuminate mechanisms of disease biology.

The *STAARpipelinePheWAS* framework supports both continuous and binary traits and uses generalized linear mixed models to adjust for population structure and relatedness^26^. Two key features drive its computational efficiency: (i) sparse genotype matrix operations, and (ii) single-pass extraction of genotypes and annotations across multiple phenotypes. Benchmarking on the UKB Research Analysis Platform (RAP) shows that per-trait analysis costs are approximately £10 across 14 masks for protein coding genes and 1 mask for ncRNA genes (**Supplementary Fig 16, Supplementary Tables 26-27**, **Supplementary Note**). While the pipeline also supports common variant analysis through single variant testing, we focused on rare variant analysis through variant set testing, given the extensive availability of GWAS for common variants^56–58^. Nonetheless, WGS provides superior resolution over array-based data, capturing nearly 19 times more variants^19^, and may uncover additional single variant associations.

Our findings highlight several diseases- and biomarker-related associations for biological and clinical relevance. For example, regulatory variants in *BCL2* were linked to lymphocytic leukemia, and ncRNA variants in *FAM30A* to chronic lymphoid leukemia, which are not captured in prior array- or exome-based analyses. Similarly, *JAK2* and *TET2* showed broad rare variant associations across hematologic traits and metabolic biomarkers. These results align with prior observations that noncoding rare variants account for a substantial portion of rare variant heritability and often reveal more trait-specific, biologically interpretable signals. Notably, we observed far more associations for biomarkers than diseases, likely due to low prevalence of many UK Biobank disease endpoints^58^.

To support community access and exploration, we developed an interactive portal (https://www.staarphewas.org/) that enables users to browse, visualize, and download all gene-trait association results. The resource allows users to query gene or phenotype-level associations across coding and noncoding masks and could accelerate hypothesis generation for functional studies and translational applications.

This study has several limitations. First, although our analyses included all available UK Biobank participants across multiple ancestries, the cohort is predominantly of European ancestry, which may limit the generalizability of rare variant associations to underrepresented populations. Second, we focused on diseases and biomarker traits; future work could apply *STAARpipelinePheWAS* to a broader spectrum of phenotypes, including accelerometer-based activity measures, imaging-derived characteristics, and detailed questionnaire data, to fully characterize the phenotypic impact of rare coding and noncoding variants. Given the current lack of large-scale WGS-based noncoding rare variant PheWAS, our findings would benefit from replication and meta-analysis of other deeply phenotyped WGS cohorts, such as All of Us and additional biobanks, to assess robustness across study designs, environments, and ancestry populations.

In summary, our study demonstrates the value of WGS studies for rare variant discovery across diseases and biomarkers and highlights the importance of including noncoding variation in association studies. The *STAARpipelinePheWAS* framework expands the analytical capabilities of phenome-wide association analysis by enabling efficient, large-scale investigation of rare coding and noncoding variants. The resulting resource, accessible through our publicly available web portal, provides an opportunity for functional interpretation, risk prediction, and therapeutic development informed by rare variant genetics.

## Methods

### UK Biobank whole genome sequencing data processing and quality control

We used the pVCF format files for WGS data of 490,549 UK Biobank participants (Data Field: 23374) and converted to annotated Genomic Data Structure (aGDS) format using *vcf2agds*^27^ for downstream analyses. Quality control followed the procedure described in a previous UK Biobank WGS study^59^. We kept all variants with pass indicated by QC label and AAScore > 0.5, where AAscore was generated by GraphTyper^60^, the software used by the UK Biobank to perform genotype calling. Variants were further filtered by excluding those that failed the Hardy-Weinberg equilibrium test (*P* < 1 × 10^−100^) or had a call rate below 90%^18,19,27^.

### Phenotypes

For disease outcomes, this study used data provided by patients and collected by the NHS as part of their routine care. We leveraged the full span of historical EHR data available in UK Biobank, integrating events from hospital inpatient records, the national death register, and the cancer registry. These events were mapped to phenotypes (phecodes) using their corresponding International Classification of Diseases (ICD) codes via the Phecode Map v1.2^61,62^. The phecode system also defines exclusion criteria for each phenotype, identifying related conditions that could indicate likely or potential undiagnosed cases of the phenotype under consideration. As an illustrative example, a previous EHR-based study of type 2 diabetes^62^ applied these exclusion rules as follows: individuals with ICD codes mapping to phecode 250.2 (“Type 2 diabetes”) were defined as cases. Controls were restricted to participants without diagnoses in the broader “Diabetes” disease group (phecodes 249-250.99), thereby avoiding contamination by conditions such as “Type 1 diabetes” (phecode 250.1).

Individuals with signs or symptoms potentially indicating undiagnosed disease, such as “Abnormal glucose” (phecode 250.4) were also excluded. In the present study, we applied analogous rules across all selected phenotypes: participants meeting any exclusion criteria for a given phecode were removed from the analysis of that phenotype to reduce the inclusion of likely prior or undiagnosed cases. Exclusions were phenotype-specific; individuals excluded from one phecode analysis were not excluded from others unless they also met the relevant criteria. Detailed exclusion rules for all phenotypes are provided in **Supplementary Table 1**.

Among the 490,549 UK Biobank participants with WGS data, we initially identified 1,695 phecodes. We retained individuals whose genetic sex was concordant with their reported sex and excluded participants with sex-specific diagnostic inconsistencies or those who had withdrawn consent. We removed the “injuries & poisonings” category because these conditions are generally not genetically mediated, and further excluded phenotypes with limited relevance to inherited risk (e.g., glossitis). To ensure adequate statistical power, we restricted analyses to phecodes with at least 500 cases in phenotype definitions. We also included proxy phenotypes based on parental or sibling disease history to define Alzheimer’s disease and related dementia^63^, as well as Parkinson’s disease (including proxy cases)^64^. After applying these criteria, 944 phecodes were included for analysis (**Supplementary Table 1**).

For quantitative traits, we first analyzed 76 blood, urine, and physical biomarkers (**Supplementary Table 2**)^15,47,48^. Blood biomarkers included Blood Count (Category 100081) and Blood Biochemistry (Category 17518), and urine biomarkers were taken from Category 100083. Four biomarkers showed highly skewed distributions and were excluded: nucleated red blood cell count, nucleated red blood cell percentage, rheumatoid factor, and microalbumin in urine. We also derived four additional phenotypes^26,47,65,66^: estimated glomerular filtration rate (eGFR), urinary albumin-to-creatinine ratio (UACR), non-albumin protein, and the Aspartate aminotransferase (AST) to Alanine aminotransferase (ALT) ratio. Additionally, for two lipid biomarkers (LDL direct, Data Field 30780, and cholesterol, Data Field 30690), we adjusted for the effects of commonly prescribed lipid-lowering medications^26^. For LDL direct, we first excluded implausible values (LDL < 10 mg/dL or Triglycerides > 400 mg/dL). Both LDL direct and cholesterol were then adjusted for statin use based on “statin adjustment factors” previously estimated, using 0.7 for LDL direct and 0.8 for cholesterol^22,24^. Triglycerides (Data Field 30870) were log-transformed before analysis.

For the plasma metabolome, concentrations of 170 blood metabolites (109 non-derived metabolites and 61 composite metabolites) and 81 metabolite ratios were measured in the phase 2 release of the UK Biobank dataset (*n* = 270*K*). These measurements were generated by Nightingale Health using nuclear magnetic resonance (NMR) spectroscopy. Metabolite values were normalized for known technical sources of variation using the *ukbnmr* R package (https://github.com/sritchie73/ukbnmr/)^48,67^. We also derived 76 additional metabolite ratios that were not included in the original Nightingale dataset but were recommended for their potential biological relevance^48,67^. In total, this yielded 327 metabolite measurements (109 non-derived metabolites, 61 composite metabolites, 81 metabolite ratios, and 76 ukbnmr-derived metabolite ratios). Five ratios showed highly skewed distributions and were excluded, resulting in 322 traits used in the final analysis. These 322 metabolites were grouped into seven major categories based on their sub-class annotations (**Supplementary Table 2**)^48^: total lipids (*n* = 17), other lipids (*n* = 6), fatty acids and apolipoproteins (*n* = 22), lipoprotein classes including their compositions (*n* = 48), lipoprotein subclasses including their compositions (*n* = 206), amino acids (*n* = 11), and others (*n* = 12). The others category included measures related to fluid balance, glucose metabolism, glycolysis intermediates, inflammation, and ketone bodies.

### Whole-genome sequencing analysis of rare coding and noncoding variants

For the 76 clinical biomarkers and 322 metabolomics traits, we first applied linear regression adjusting for age, age^2^, sex, and the first 10 ancestral principal components (PCs). For the metabolomics traits, BMI was additionally included, and statin use (binary variable) was further adjusted for the 299 lipid-related measurements^48^. The residuals were then rank-based inverse normal transformed and rescaled using the standard deviation of the original phenotype. We next fitted a linear mixed model (LMM) on the normalized residuals using the same covariates, incorporating a variance component for an empirically-derived sparse genetic relatedness matrix (GRM) using *FastSparseGRM*^27,68^ to account for population structure and sample relatedness. For the 944 diseases, we applied logistic mixed models with identical covariate adjustments and GRM specifications.

Variant set analysis was performed on aggregated rare autosomal variants (minor allele frequency [MAF] < 1%)^26,27^. For the 76 clinical biomarkers and 322 metabolomics traits, we computed the *P* value of each variant set using STAAR-O^26^, an omnibus test aggregating burden test^69^, SKAT^70^, and ACAT-V^71^ in the STAAR framework^72^. For the 944 diseases, we calculated the *P* value of each variant set using STAAR-Burden. *P* values were initially computed using the normal approximation, and when the resulting *P* value was < 0.05, we recalibrated it using the saddlepoint approximation (SPA)^73^ to improve accuracy. The gene-centric coding analysis of variants, including both single-nucleotide variants (SNVs) and indels, provides 7 coding functional categories of protein coding genes, including putative loss of function (stop gain, stop loss and splice) variants, missense variants, disruptive missense variants, putative loss of function and disruptive missense variants, synonymous variants, protein-truncating RVs (stop gain, stop loss, splice, frameshift deletion and frameshift insertion), and protein-truncating RVs and disruptive missense RVs. The putative loss of function, missense, synonymous, and protein-truncating RVs were defined by GENCODE Variant Effect Predictor (VEP) categories^74,75^. The disruptive variants were further defined by MetaSVM^76^, which measures the deleteriousness of missense mutations.

The gene-centric noncoding analysis provides 8 genetic categories of variants, including promoter or enhancer overlaid with CAGE or DHS sites, UTR, upstream, downstream of protein coding genes, and noncoding RNA genes. The promoter RVs are defined as RVs in the +/− 3-kilobase (kb) window of transcription start sites with the overlap of CAGE sites or DHS sites. The enhancer RVs are defined as RVs in GeneHancer predicted regions with the overlap of CAGE sites or DHS sites^77–80^. We define the UTR, upstream, downstream, and ncRNA RVs by GENCODE VEP categories^74,75^. For the UTR mask, we include RVs in both 5’ and 3’ UTR regions. For the ncRNA mask, we include the exonic and splicing ncRNA RVs. We consider the protein-coding genes for the first seven categories provided by Ensembl^81^ and the ncRNA genes provided by GENCODE^74,75^.

### STAARpipelinePheWAS framework

To analyze a broad spectrum of phenotypes from large-scale whole-genome sequencing datasets such as the UK Biobank^19^, All of Us^34^, and TOPMed^82^, we developed *STAARpipelinePheWAS* (STAARpipeline for Phenome-Wide Association Analysis). This framework enables efficient and scalable association analyses of both coding and noncoding variants. By leveraging the inherent sparsity of rare variant genotype data, *STAARpipelinePheWAS* extracts and operates on sparse genotype matrices throughout the entire workflow, reducing memory use and computation time without loss of information. Genotype data and functional annotations are extracted only once for multiple phenotypes, which allows high-throughput rare-variant analyses at biobank scale. Benchmarking in the UK Biobank Research Analysis Platform (UKB-RAP) showed more than a 4-fold cost reduction on average, compared to analyzing each phenotype separately (**Supplementary Note**).

### Drug target validation

We evaluated the enrichment of drug targets among the identified protein-coding genes from coding analyses using four publicly accessible resources: DrugBank (https://www.drugbank.ca/)^38^, the Therapeutic Target Database (TTD; https://db.idrblab.net/ttd/)^83^, the Informa Pharmaprojects database (https://raw.githubusercontent.com/AbbVie-ComputationalGenomics/genetic-evidence-approval/master/data/target_indication.tsv)^15^, and curated druggable genome sets^84^.

For DrugBank, we included genes annotated as “Approved” or “Approved | Investigational” (*n* = 1,750). For TTD, we included genes labeled as “Clinical trial” or “Successful” (*n* = 1,476). For Informa Pharmaprojects, we included genes categorized as “Approved”, “Phase I Clinical Trial”, “Phase II Clinical Trial”, or “Phase III Clinical Trial” (*n* = 1,267). The druggable genome reference set consisted of 4,182 protein-coding genes.

To assess enrichment, we applied Fisher’s exact test to evaluate the relationship between drug target status and the identified protein-coding genes. For each of the four drug target gene lists, we created a contingency table that included the number of protein-coding genes identified in coding analyses that intersected with the list and the number of genes that did not intersect with the list out of the 18,445 genes tested in the PheWAS^15^.

### Protein-protein interaction analysis

We used the Search Tool for the Retrieval of Interacting Genes database (STRING) database (Version 12.0, https://string-db.org/)^85^ to investigate protein-protein interactions (PPIs) among the identified protein-coding genes from both coding and noncoding analyses. Interactions supported by experimental evidence, curated database entries, or co-expression were retained, and interaction pairs with a combined score greater than 0.4 (medium confidence) were considered significant^86^. Gene clusters were identified using a K-means clustering approach. The default number of clusters (*K*) was set to 3. When STRING reported that this value was smaller than the number of disconnected components in the network, the software automatically increased *K* to ensure coherent clustering. This adjustment allowed the clusters to match the natural structure of the PPI network.

### Functional enrichment analysis

To interpret the biological pathways represented by the identified protein-coding genes from both coding and noncoding analyses, we used STRING to perform functional enrichment across three widely used annotation resources^85^: Gene Ontology (GO) Biological Process^87^, the Kyoto Encyclopedia of Genes and Genomes (KEGG)^88^, and Reactome^89^. For each gene set, the top ten significantly enriched pathways from each analysis (GO, KEGG, and Reactome) are shown. Pathways with a false discovery rate less than 0.05 were considered statistically significant.

### Overlap with the GWAS Catalog

To assess supporting evidence from prior GWAS, we systematically queried the NHGRI-EBI GWAS Catalog (downloaded on 5 December 2025)^28^. Disease traits were mapped using phecodes or disease names, while biomarkers were matched using UK Biobank Field IDs or trait names. We retained genome-wide significant associations (*P* < 5 × 10^−8^) with available genomic coordinates. A gene identified in our coding or noncoding rare variant analyses was considered supported by previous GWAS if it was located within 500 kilobase (kb) of a reported GWAS variant for the corresponding trait^19,90,91^.

### Overlap with prior sequencing-based studies

To assess the extent to which our coding and noncoding associations overlapped with previous sequencing-based analyses, we compared our WGS findings for 944 diseases and 76 clinical biomarkers with results from the AstraZeneca PheWAS portal (https://azphewas.com)^15,19^, a large repository of gene-phenotype associations derived from electronic health records, questionnaire data, and continuous traits in UK Biobank exome and genome data using gene-based collapsing analysis. AstraZeneca defines disease phenotypes primarily using UK Biobank tree fields, such as ICD-10 hospital admissions (field 41202)^15^, whereas our analyses rely on phecodes^61,62^. To ensure a fair comparison, we constructed a phecode-ICD-10 correspondence table (**Supplementary Table 1**) and queried the AstraZeneca portal using all ICD-10 codes mapped to each phecode. Because a single phecode may correspond to multiple ICD-10 codes, for example, phecode 153 (Colorectal cancer) maps to C18, C19, C20, C210, C211, C212, C218, C260, D010, D011, D012, and D013, we searched all corresponding codes and retained only ICD-10 phenotypes with at least 200 cases^19^. When multiple related AstraZeneca phenotypes were available for a given ICD-10 code, we prioritized the “union” phenotypes, which aggregate cases across closely related disease definitions to improve comparability.

For the 322 plasma metabolomic traits, we compared our WGS findings with three sequencing-based resources: the AstraZeneca PheWAS portal (phase 1)^19,48^, the phase 2 dataset^50^, and the phase 3 dataset^51^.

## Genome build

All genome coordinates are given in NCBI GRCh38/UCSC hg38.

## Data availability

Association statistics generated in this study are publicly available through our PheWAS Portal (https://www.staarphewas.org/). All whole-genome sequencing data described in this paper are publicly available to registered researchers through the UKB data access protocol. The genome data can be found via the UK Biobank Showcase (Data Field: 23374): https://biobank.ndph.ox.ac.uk/ukb/field.cgi?id=23374. Information on how to apply for data access is available at https://www.ukbiobank.ac.uk/use-our-data/apply-for-access/. Data for this study were obtained under UK Biobank applications 91486, 52008 and 211447.

## Code availability

*STAARpipelinePheWAS* is implemented as an open-source R package available at https://github.com/li-lab-genetics/STAARpipelinePheWAS, and as an applet in UK Biobank RAP available at https://github.com/li-lab-genetics/STAARpipelinePheWAS-rap-batch. *STAARpipelineSummary* is available at https://github.com/xihaoli/STAARpipelineSummary and as a RAP applet available at https://github.com/li-lab-genetics/staarpipelinesummary_varset-rap. The *vcf2agds* toolkit (https://github.com/drarwood/vcf2agds_overview) was used to preprocess UK Biobank WGS data. *FastSparseGRM* (https://github.com/rounakdey/FastSparseGRM) was used to construct empirically-derived sparse genetic relatedness matrix (GRM) and ancestral principal components (PCs). Software development and construction of coding and noncoding analyses for UK Biobank WGS data were performed under UK Biobank applications 91486, 52008 and 211447. All analyses were conducted on the UK Biobank RAP (https://ukbiobank.dnanexus.com/). The *createUKBphenome* package (https://github.com/umich-cphds/createUKBphenome) was used to extract and map ICD-coded hospital records to phecodes for harmonized case/control phenotypes. The STRING v12.0 database (https://string-db.org/) was used for protein-protein interaction and functional enrichment analyses.

## Acknowledgements

We thank the participants and investigators of the UK Biobank for making this research possible. X. Lin is supported by NIH grants R35-CA197449, R01-HL163560, U19-CA203654, U01-HG012064, and U01-HG009088. We also thank Biomarker Technologies for supporting the development of the PheWAS Portal.

## Author contributions

Xihao Li, X. Lin, and Z. Li designed the experiments. Y. Yuan, Xihao Li, X. Lin, and Z. Li performed the experiments. Y. Yuan, Y. Guan, Y. Feng, T. Chen, Y. Zhang, B. Chang, S. Fan, C. Lu, W. Li, Xiaoyu Li, Xihao Li, X. Lin, and Z. Li acquired, analyzed, or interpreted data. Y. Yuan, Xihao Li and Z. Li developed and maintained the software. Y. Yuan, Xihao Li, X. Lin, and Z. Li drafted the manuscript and revised it according to co-authors’ suggestions. All authors critically reviewed the manuscript, suggested revisions as needed, and approved the final version.

