## Supplementary Information for "Rare coding and noncoding variants map 1,342 diseases and biomarkers in 490,549 whole genomes"

#### Supplementary Note

##### Biological insights into the protein-coding genes identified from coding and noncoding analyses for 398 biomarkers

The 398 quantitative biomarkers were grouped into 11 major categories based on their subclass annotations (**Supplementary Table 2**)[1]: blood count ($n = 29$), blood biochemistry ($n = 32$), urine assays ($n = 4$), physical measurements ($n = 11$), NMR total lipids ($n = 17$), NMR other lipids ($n = 6$), NMR fatty acids and apolipoproteins ($n = 22$), NMR lipoprotein classes including their compositions ($n = 48$), NMR lipoprotein subclasses including their compositions ($n = 206$), NMR amino acids ($n = 11$), and NMR others ($n = 12$). To gain biological insight into these biomarker-associated genes, we used the Search Tool for the Retrieval of Interacting Genes database (STRING) database (Version 12.0, https://string-db.org/)[2] to evaluate protein-protein interactions (PPIs) and conduct functional enrichment analyses. Enrichment was performed across three widely used annotation resources: GO Biological Process[3], KEGG[4] and Reactome[5]. Because STRING supports a maximum of 2,000 proteins per query, we applied a stringent threshold of $\alpha = 1.00 \times 10^{-8}$ for selecting protein-coding genes in the blood count and blood biochemistry categories[1,6,7]. For the remaining nine categories, we applied a Bonferroni-corrected genome-wide significance threshold of

$\alpha = 0.05/(20{,}000 \times 7) = 3.57 \times 10^{-7}$, accounting for 7 different coding or noncoding masks across protein-coding genes. In total, the blood count, blood biochemistry, urine assays, physical measurements, NMR total lipids, NMR other lipids, NMR fatty acids and apolipoproteins, NMR lipoprotein classes and their components, NMR lipoprotein subclasses and their components, NMR amino acids, and NMR other traits categories identified 1,538; 1,573; 37; 663; 259; 208; 719; 386; 1,101; 231; and 251 protein-coding genes, respectively.

PPI network analyses revealed that most protein-coding genes within each biomarker category clustered into a single dominant module, indicating convergence on coherent biological pathways (**Supplementary Figs 5-15a**). Functional enrichment using GO Biological Process, KEGG, and Reactome further highlighted biologically meaningful patterns across traits. Specifically, genes for blood count traits were enriched in hematopoiesis, myeloid and lymphoid differentiation, granulopoiesis, and immune processes such as megakaryocyte development and antigen processing, reflecting core pathways of blood cell formation (**Supplementary Fig 5b**, **Supplementary Table 15**). Blood biochemistry genes were enriched in lipid and cholesterol homeostasis, plasma lipoprotein remodeling, and triglyceride metabolism (**Supplementary Fig 6b**, **Supplementary Table 16**), whereas urine biomarker genes mapped to glomerular basement membrane development, collagen crosslinking, and interferon-gamma signaling, consistent with renal and immune functions (**Supplementary Fig 7b**, **Supplementary Table 17**). Genes for physical measurement traits were enriched in chondrocyte differentiation, skeletal development, ossification, extracellular matrix organization, and hormone response pathways (**Supplementary Fig 8b**, **Supplementary Table 18**). For NMR-based metabolomics, which predominantly captures lipid-related traits[8], lipid-associated genes were enriched in triglyceride, cholesterol, and lipid homeostasis and in the regulation of plasma lipoprotein particles (**Supplementary Figs 9-13b**, **Supplementary Tables 19-23**). Genes for NMR amino acid traits were enriched in amino acid catabolism, small molecule metabolism, branched-chain amino acid degradation, and transport (**Supplementary Fig 14b**, **Supplementary Table 24**). NMR other traits, including fluid balance, glucose metabolism, glycolysis, inflammation, and ketone bodies, showed enrichment in triglyceride and phospholipid metabolism, glucagon and PPAR signaling, glycolysis/gluconeogenesis, and insulin signaling (**Supplementary Fig 15b**, **Supplementary Table 25**). Together, these results demonstrate that the identified gene-trait associations map consistently onto coherent and biologically interpretable pathways across a diverse set of biomarkers.

**Overview of the methods**

To analyze this wide range of phenotypes in 490,549 UK Biobank genomes, we first leverage the inherent sparsity of rare variant genotypes. By directly extracting and operating on sparse genotype matrices throughout the STAAR series pipeline (incorporating updates from *STAARpipeline* versions 0.9.8 to 0.9.9), both memory usage and computation time are greatly reduced without loss of information, enabling efficient biobank-scale WGS/WES analyses. For example, *TTN* includes ~23,000 rare missense variants: extracting a dense genotype matrix requires 78 GB of memory in

STAARpipeline v0.9.8, whereas sparse extraction in *STAARpipeline* v0.9.9 requires only 47 MB.

Phenome-wide analyses typically require repeated extraction of genotype matrices and functional annotations for each trait, which becomes increasingly costly when hundreds or thousands of phenotypes are analyzed. To address this, we developed *STAARpipelinePheWAS* (STAARpipeline for Phenome-Wide Association Analysis), which implements a single-pass extraction procedure that retrieves genotypes and annotations once and reuses them across all traits (**Fig. 1b**). This design markedly reduces computational burden and substantially improves scalability for WGS-based PheWAS.

To further control computational costs when using spot instances on cloud platforms such as the UKB RAP, we partitioned chromosome 1-22 analyses into smaller jobs. For example, gene-centric coding analyses with 7 masks were divided into 164 jobs, gene-centric noncoding analyses with 7 masks were also divided into 164 jobs, and ncRNA analyses were divided into 65 jobs. This strategy ensured that most jobs finished within one hour. Both *STAARpipeline* and *STAARpipelinePheWAS* support execution on cloud environments and high-performance computing clusters.

We benchmarked computation time (in hours) and cloud cost (£) for *STAARpipeline* v0.9.8, *STAARpipeline* v0.9.9, and *STAARpipelinePheWAS* v0.9.7.1 using UK Biobank 500K WGS data for rare coding and noncoding variant set analyses (**Supplementary**

**Fig 16**, **Supplementary Tables 26-27**). *STAARpipeline* v0.9.9 reduced cloud cost by 2.93-fold for quantitative traits and 3.49-fold for imbalanced case-control traits compared with *STAARpipeline* v0.9.8. For noncoding analyses (7 masks), the reductions were 4.21-fold and 5.01-fold, respectively. These improvements come mainly from leveraging the inherent sparsity of rare variant genotype data, as implemented in *STAARpipelinePheWAS*.

Furthermore, when analyzing 10 phenotypes simultaneously, *STAARpipelinePheWAS* achieved 3.97-fold cost reductions for quantitative traits and 4.62-fold reductions for imbalanced case-control traits compared to *STAARpipeline* v0.9.9. In addition to operating directly on sparse genotype matrices, the pipeline retrieves genotypes and functional annotations once and reuses them across all phenotypes, enabling high-throughput coding and noncoding rare variant analyses at biobank scale. Taken together, these design features provide more than a 4-fold improvement in computational efficiency relative to *STAARpipeline* v0.9.9, which analyzes each phenotype separately.

### Supplementary Figures

**Supplementary Figure 1 | Distribution and sample sizes of the diseases and biomarkers. a** Percentage of diseases included in the study, grouped by Phecode-based category. **b** Median number of cases for each disease across categories. Bars indicate the interquartile range (IQR). **c** Percentage of health-related biomarkers included in the study. **d** Median sample size per biomarker across categories. Bars indicate the interquartile range (IQR).

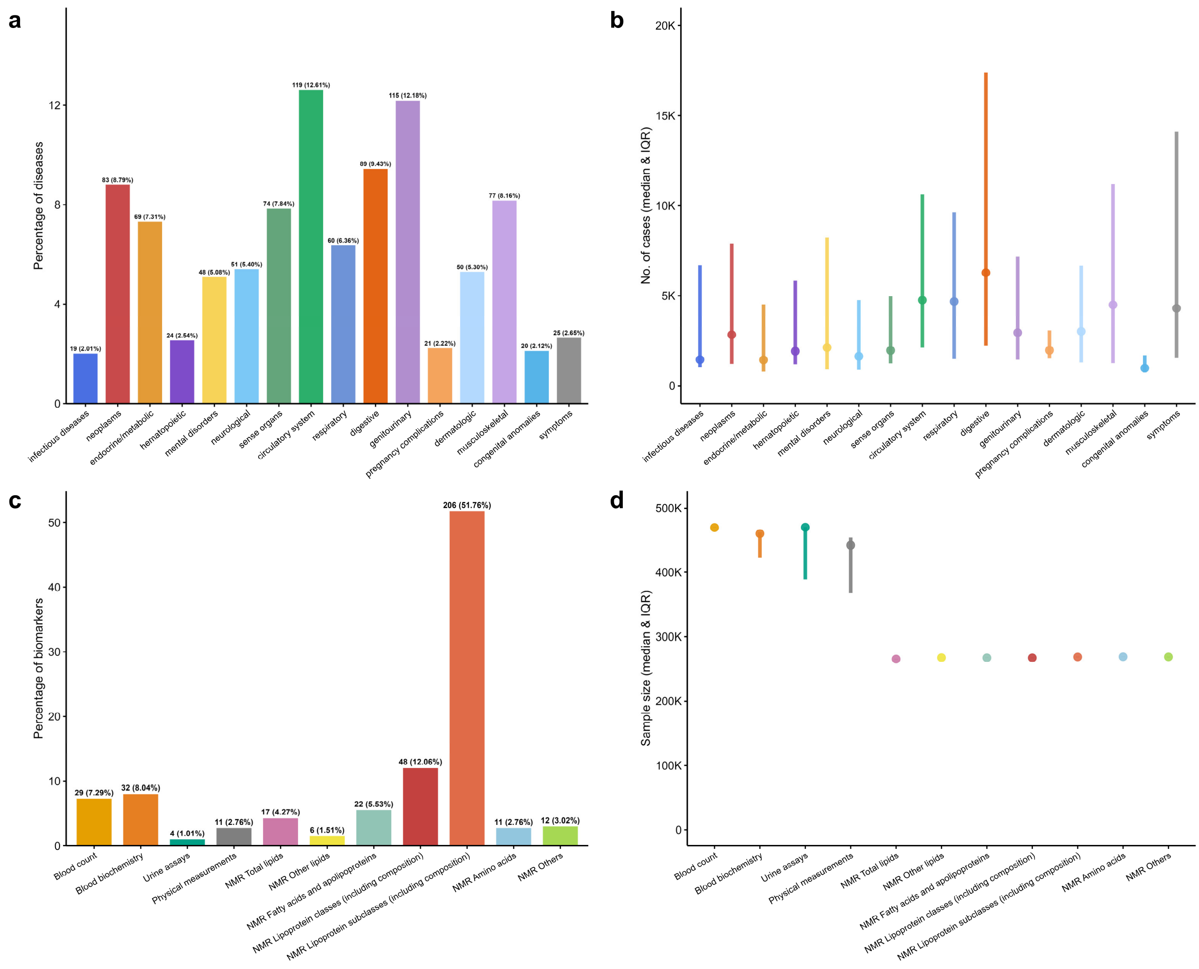

**Supplementary Figure 2 | Summary of coding analyses (7 masks) and noncoding analyses (8 masks) for 398 biomarkers. a** Gene-centric coding analyses (7 masks) for biomarkers, colored by biomarker category. **b** Frequency of mask-trait associations across 7 coding masks grouped by biomarker category. **c** Gene-centric noncoding analyses (8 masks) for biomarkers, colored by biomarker category. **d** Frequency of mask-trait associations across 8 noncoding masks grouped by biomarker category. In panels **a** and **c**, each column on the x-axis represents a trait, and each point represents a mask-trait association. The dashed line indicates the genome-wide significance threshold ($\alpha = 0.05/(20{,}000 \times 7) = 3.57 \times 10^{-7}$). The y-axis is capped at $-\log_{10}(P) = 300$, and associations with $P < 0.01$ are shown.

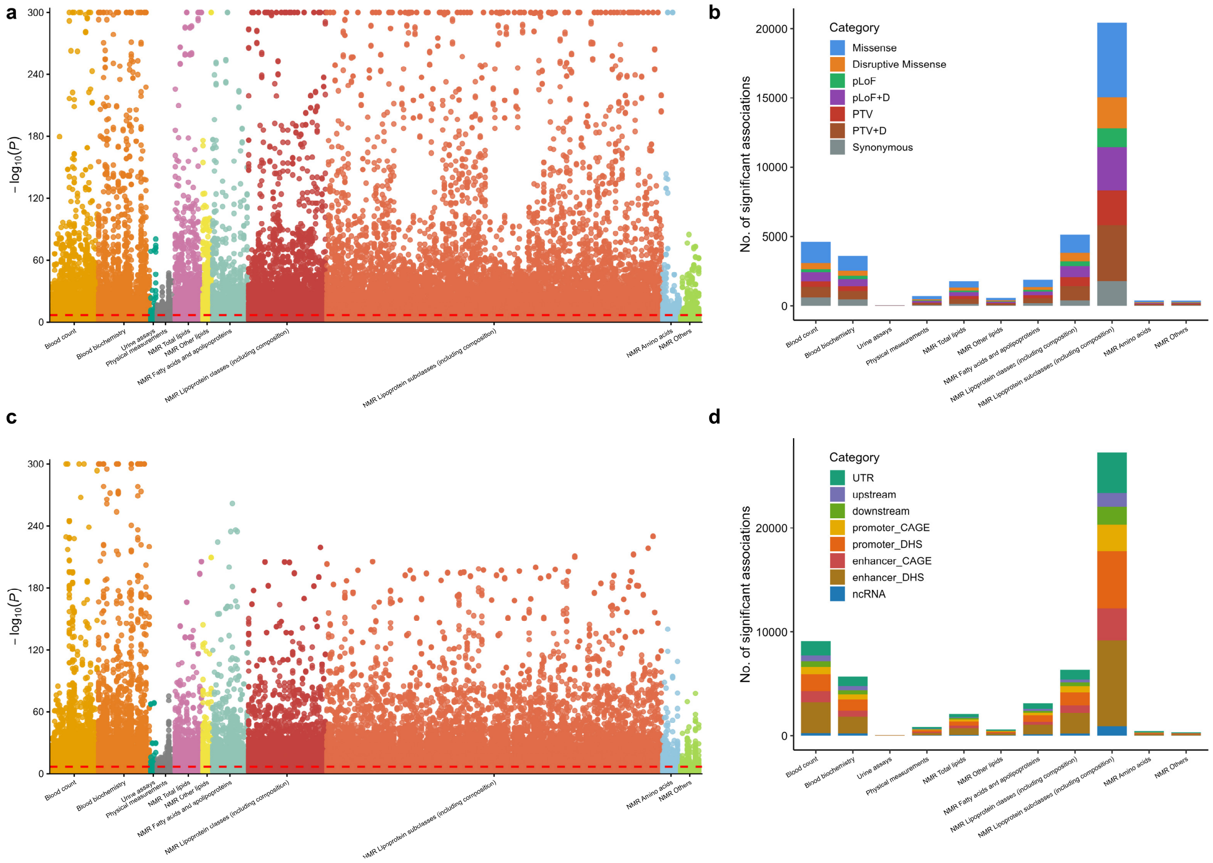

**Supplementary Figure 3 |** Enrichment of FDA-approved drug targets among protein-coding genes identified from coding analyses. Error bars represent 95% CIs. Pharmaprojects and Druggable Genome were used as drug-target reference databases. $P$ values were calculated using a two-sided Fisher's exact test. The first row represents enrichment across all 398 biomarkers.

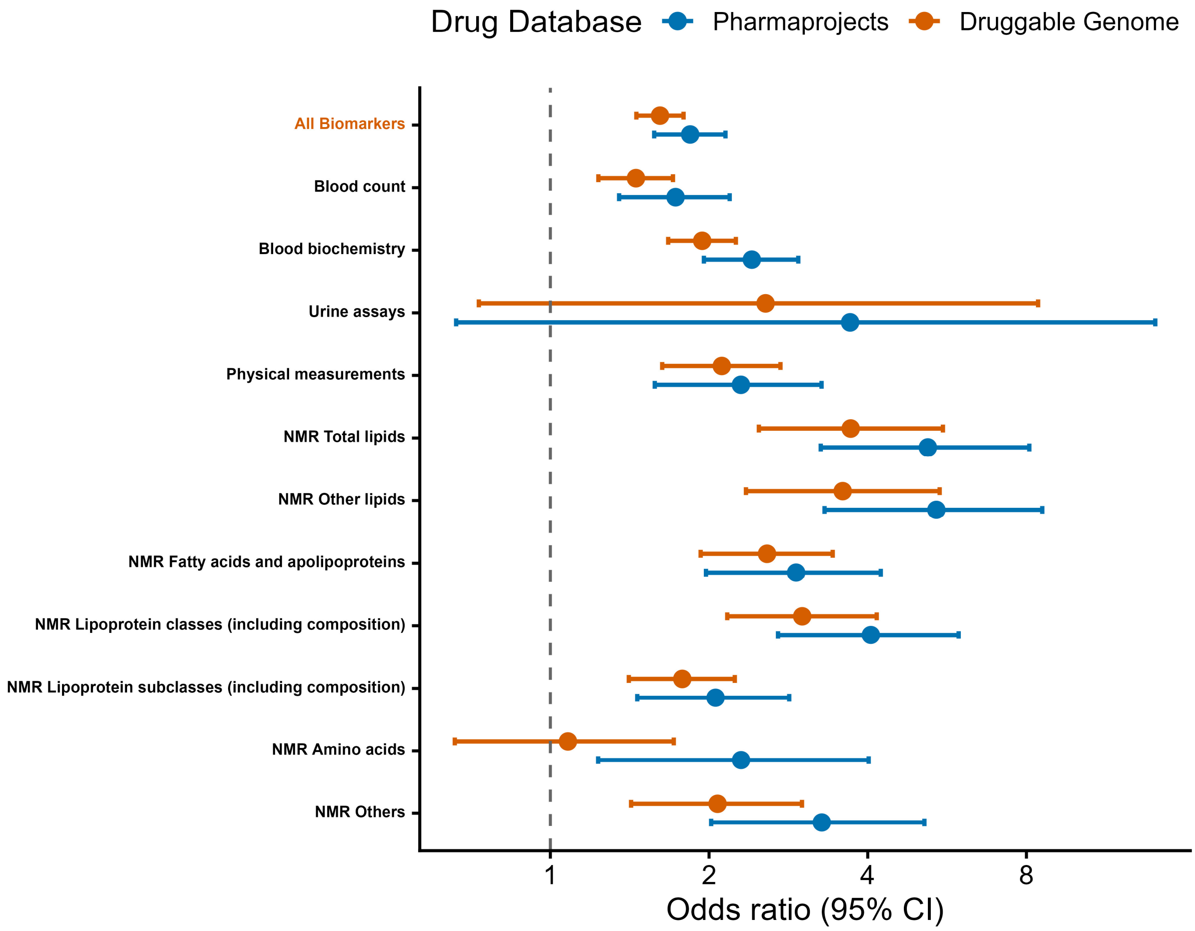

**Supplementary Figure 4 | Summary of rare coding and noncoding genetic variants associated with 398 biomarkers.** Top row (**a**-**c**): coding rare variant analyses. Bottom row (**d**-**f**): noncoding rare variant analyses. **a** Top three biomarkers within each biomarker category showing the largest numbers of significant gene-trait pairs identified in coding analyses. Bars are colored by biomarker category, and gene labels correspond to the most significant association (minimum $P$ value) for each biomarker. Biomarkers with at least three significant gene associations are included. Mean corpuscular haemoglobin (MCH), MCH; Mean platelet (thrombocyte) volume (MPV), MPV; Mean corpuscular volume (MCV), MCV; Alkaline phosphatase (ALP), ALP; urinary albumin-to-creatinine ratio, UACR; Creatinine (enzymatic) in urine, Creatinine; Sodium in urine, Sodium; Forced vital capacity (FVC, best measure), FVC; Pulse rate (automated reading), Pulse rate. See Supplementary Table 2 for abbreviations of NMR metabolites. **b** Significant gene-trait pairs from coding analyses for two representative biomarkers in each biomarker category. For biomarkers with multiple mask-specific associations, the minimum $P$ value per gene is shown. Genes with $P < 1 \times 10^{-80}$ are labeled. **c** Stacked bar chart summarizing biomarker roles of protein-coding genes identified in coding analyses, grouped by biomarker category. Shown are the top 50 genes with the highest numbers of significant gene-trait associations; numbers above bars denote the total number of associated biomarkers. **d** Top three biomarkers within each biomarker category showing the largest numbers of significant gene-trait pairs identified in noncoding analyses. Bars are colored by biomarker category, and gene labels correspond to the most significant association (minimum $P$ value) for each biomarker. Biomarkers with at least three significant gene associations are included. Mean corpuscular haemoglobin (MCH), MCH; Mean platelet (thrombocyte) volume (MPV), MPV; Mean corpuscular volume (MCV), MCV; Alkaline phosphatase (ALP), ALP; Glycated haemoglobin (HbA1c), HbA1c; Creatinine (enzymatic) in urine, Creatinine; Sodium in urine, Sodium; urinary albumin-to-creatinine ratio, UACR; Forced vital capacity (FVC, best measure), FVC; FEV1/FVC ratio Z-score, FEV1/FVC. See Supplementary Table 2 for abbreviations of NMR metabolites. **e** Significant gene-trait pairs from noncoding analyses for two representative biomarkers in each biomarker category. For biomarkers with multiple mask-specific associations, the minimum $P$ value per gene is shown. Genes with $P < 1 \times 10^{-80}$ are labeled. **f** Stacked bar chart summarizing biomarker roles of protein-coding genes identified in noncoding analyses, grouped by biomarker category. Shown are the top 50 genes with the highest numbers of significant gene-trait associations; numbers above bars denote the total number of associated biomarkers.

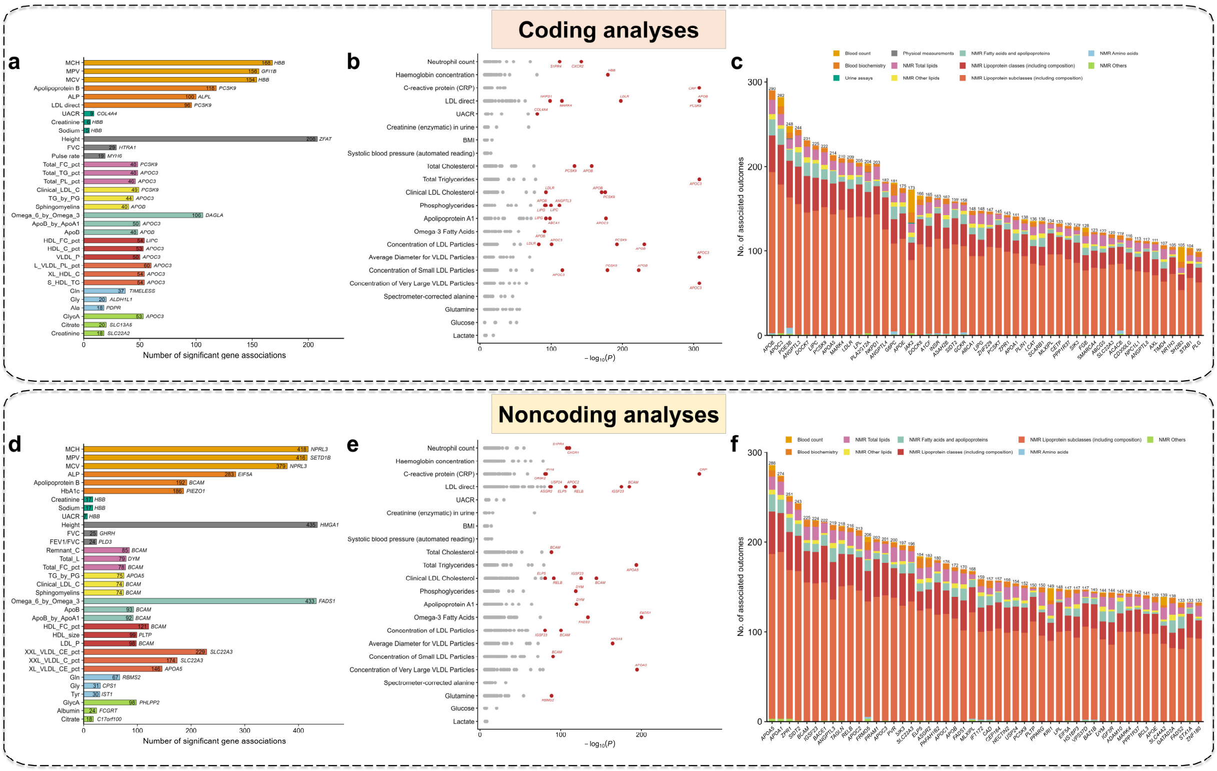

**Supplementary Figure 5 | Protein-protein interaction network and functional enrichment analyses of protein-coding genes identified from coding and noncoding analyses associated with blood count traits.** We applied Bonferroni-corrected genome-wide significance threshold of $\alpha = 1.00 \times 10^{-8}$ for protein-coding genes. Orphan genes (i.e., genes without connections in the PPI network) were hidden. **a** PPI network constructed from the identified protein-coding genes. **b** Significantly enriched pathways identified through Gene Ontology (GO), KEGG, and Reactome analyses, displaying the top four enriched pathways from each database.

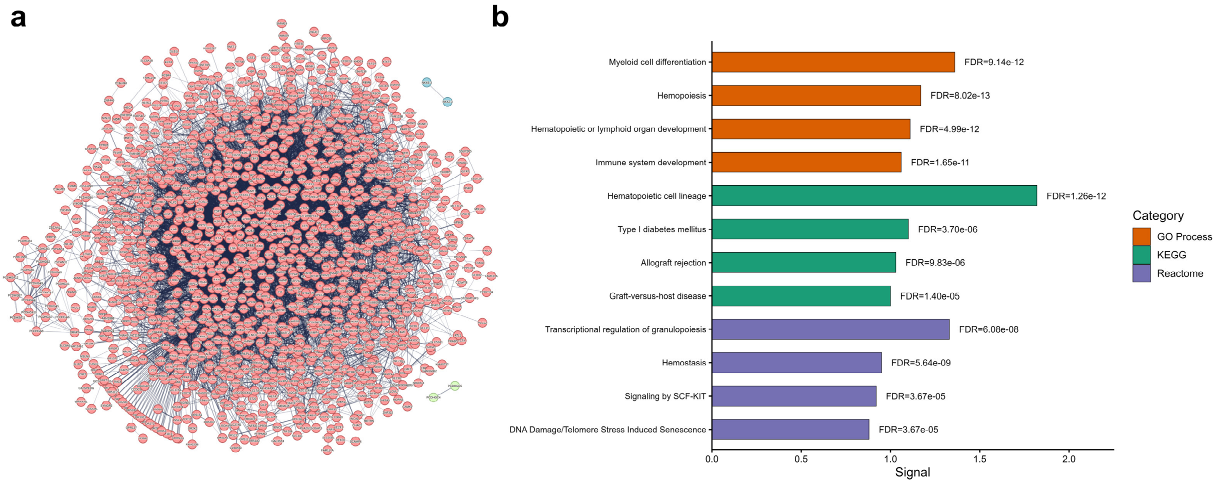

**Supplementary Figure 6 | Protein-protein interaction network and functional enrichment analyses of protein-coding genes identified from coding and noncoding analyses associated with blood biochemistry traits.** We applied Bonferroni-corrected genome-wide significance threshold of $\alpha = 1.00 \times 10^{-8}$ for protein-coding genes. Orphan genes (i.e., genes without connections in the PPI network) were hidden. **a** PPI network constructed from the identified protein-coding genes. **b** Significantly enriched pathways identified through Gene Ontology (GO), KEGG, and Reactome analyses, displaying the top four enriched pathways from each database.

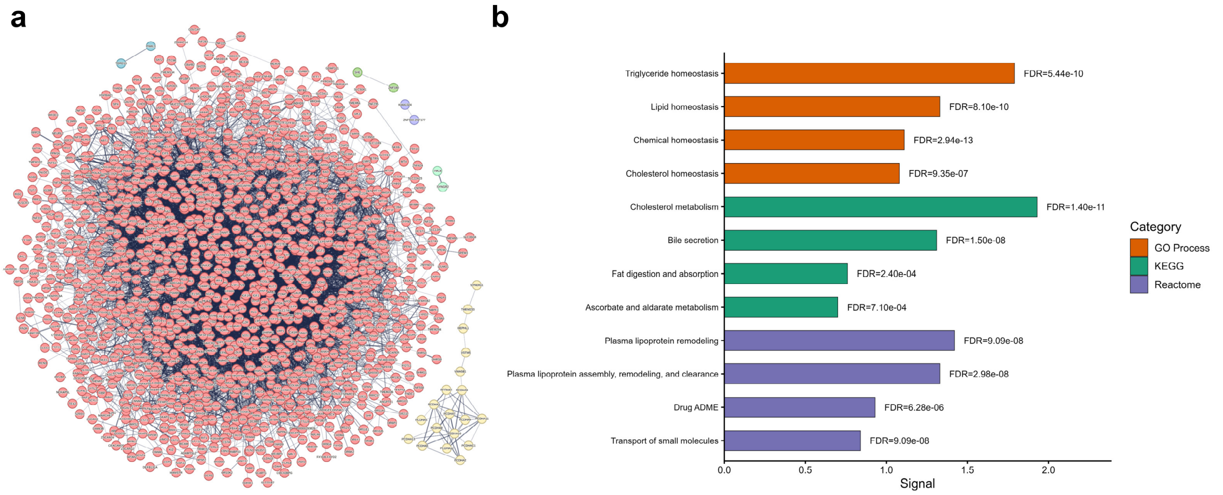

**Supplementary Figure 7 | Protein-protein interaction network and functional enrichment analyses of protein-coding genes identified from coding and noncoding analyses associated with urine assays traits.** We applied Bonferroni-corrected genome-wide significance threshold of $\alpha = 0.05/(20{,}000 \times 7) = 3.57 \times 10^{-7}$ for protein-coding genes. Orphan genes (i.e., genes without connections in the PPI network) were hidden. **a** PPI network constructed from the identified protein-coding genes. **b** Significantly enriched pathways identified through Gene Ontology (GO), KEGG, and Reactome analyses, displaying the top four enriched pathways from each database.

**a** **b**

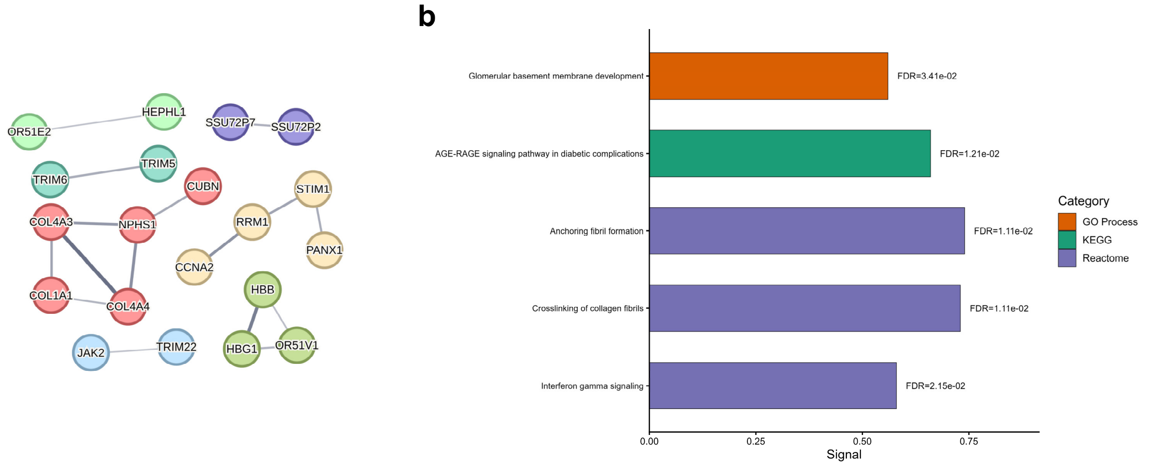

**Supplementary Figure 8 | Protein-protein interaction network and functional enrichment analyses of protein-coding genes identified from coding and noncoding analyses associated with physical measurements traits.** We applied Bonferroni-corrected genome-wide significance threshold of $\alpha = 0.05/(20{,}000 \times 7) = 3.57 \times 10^{-7}$ for protein-coding genes. Orphan genes (i.e., genes without connections in the PPI network) were hidden. **a** PPI network constructed from the identified protein-coding genes. **b** Significantly enriched pathways identified through Gene Ontology (GO), KEGG, and Reactome analyses, displaying the top four enriched pathways from each database.

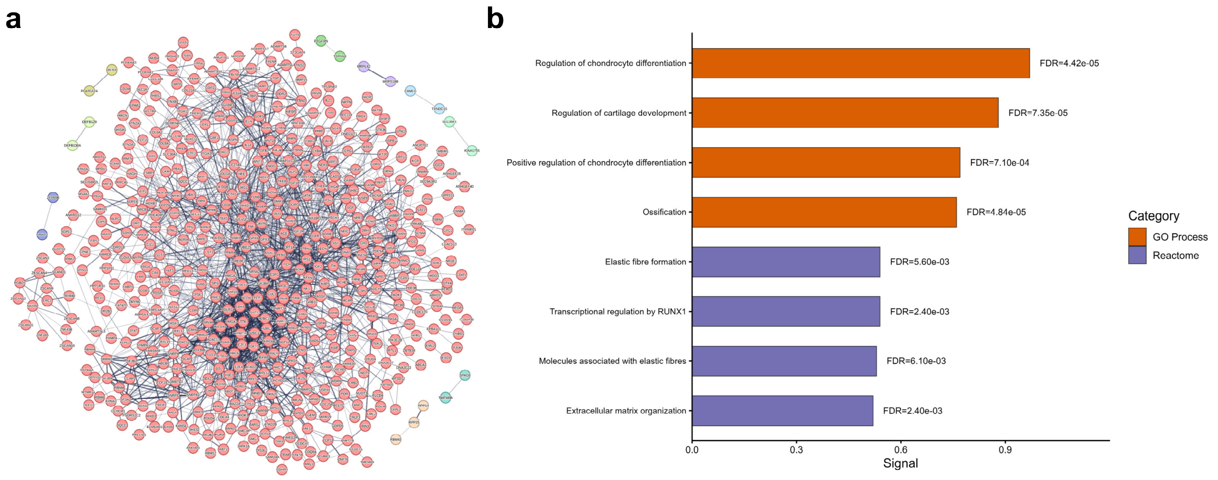

**Supplementary Figure 9 | Protein-protein interaction network and functional enrichment analyses of protein-coding genes identified from coding and noncoding analyses associated with NMR Total lipids traits.** We applied Bonferroni-corrected genome-wide significance threshold of $\alpha = 0.05/(20{,}000 \times 7) = 3.57 \times 10^{-7}$ for protein-coding genes. Orphan genes (i.e., genes without connections in the PPI network) were hidden. **a** PPI network constructed from the identified protein-coding genes. **b** Significantly enriched pathways identified through Gene Ontology (GO), KEGG, and Reactome analyses, displaying the top four enriched pathways from each database.

**a** **b**

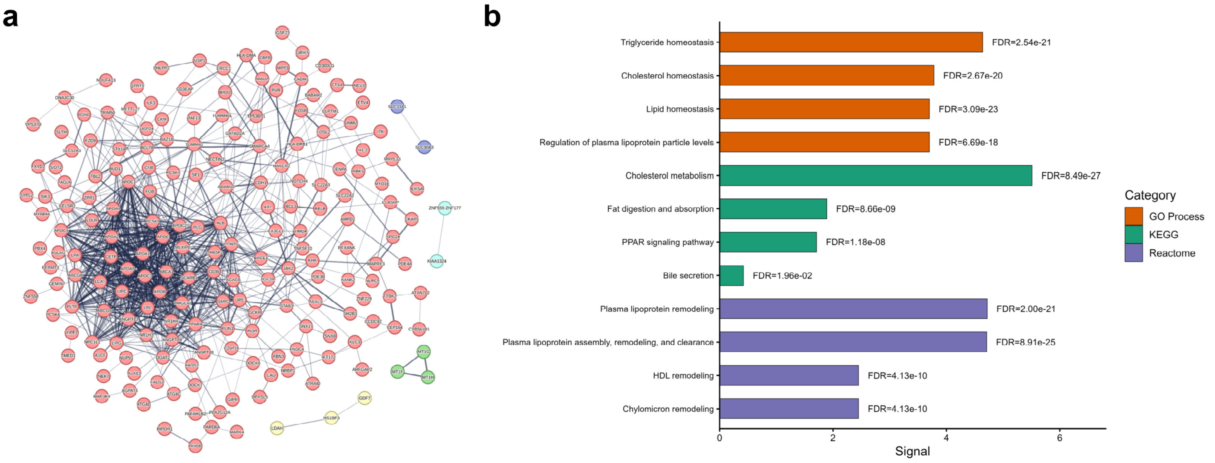

**Supplementary Figure 10 | Protein-protein interaction network and functional enrichment analyses of protein-coding genes identified from coding and noncoding analyses associated with NMR Other lipids traits.** We applied Bonferroni-corrected genome-wide significance threshold of $\alpha = 0.05/(20{,}000 \times 7) = 3.57 \times 10^{-7}$ for protein-coding genes. Orphan genes (i.e., genes without connections in the PPI network) were hidden. **a** PPI network constructed from the identified protein-coding genes. **b** Significantly enriched pathways identified through Gene Ontology (GO), KEGG, and Reactome analyses, displaying the top four enriched pathways from each database.

**a** **b**

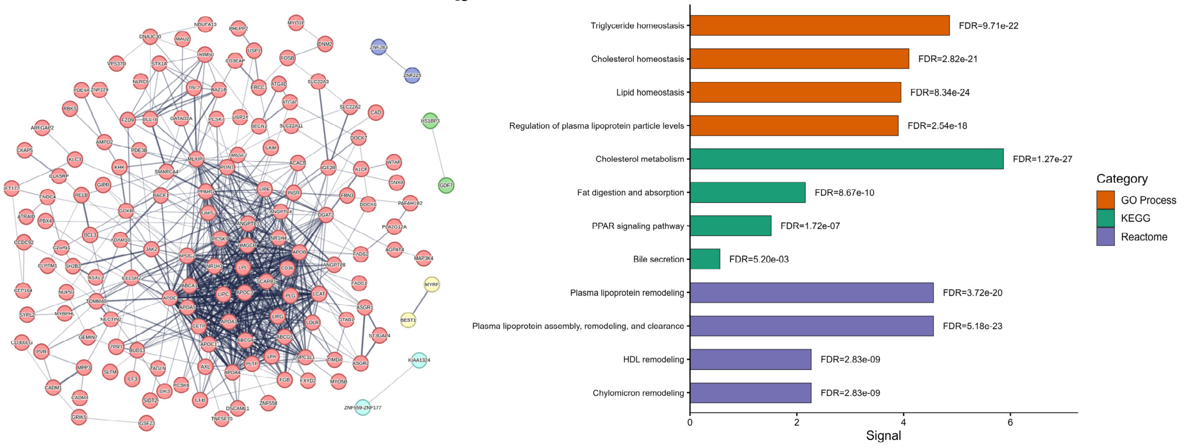

**Supplementary Figure 11 | Protein-protein interaction network and functional enrichment analyses of protein-coding genes identified from coding and noncoding analyses associated with NMR Fatty acids and apolipoproteins traits.** We applied Bonferroni-corrected genome-wide significance threshold of $\alpha = 0.05/(20{,}000 \times 7) = 3.57 \times 10^{-7}$ for protein-coding genes. Orphan genes (i.e., genes without connections in the PPI network) were hidden. **a** PPI network constructed from the identified protein-coding genes. **b** Significantly enriched pathways identified through Gene Ontology (GO), KEGG, and Reactome analyses, displaying the top four enriched pathways from each database.

**a**

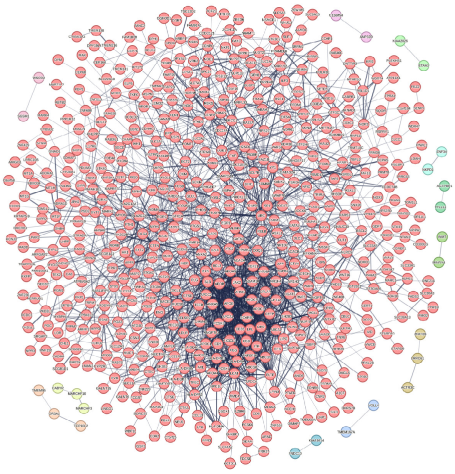

**b**

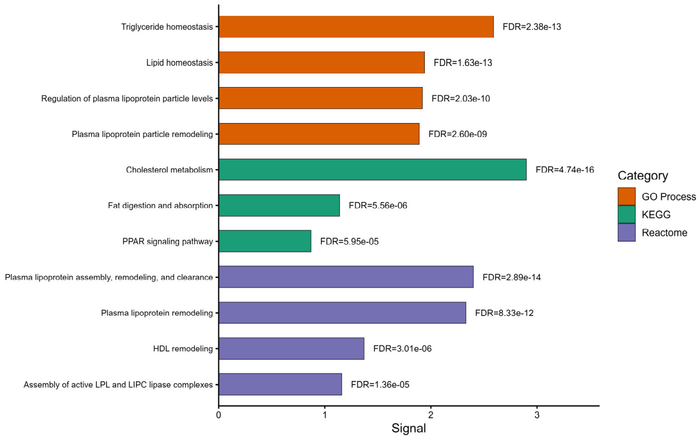

**Supplementary Figure 12 | Protein-protein interaction network and functional enrichment analyses of protein-coding genes identified from coding and noncoding analyses associated with NMR Lipoprotein classes (including composition) traits.** We applied Bonferroni-corrected genome-wide significance threshold of $\alpha = 0.05/(20{,}000 \times 7) = 3.57 \times 10^{-7}$ for protein-coding genes. Orphan genes (i.e., genes without connections in the PPI network) were hidden. **a** PPI network constructed from the identified protein-coding genes. **b** Significantly enriched pathways identified through Gene Ontology (GO), KEGG, and Reactome analyses, displaying the top four enriched pathways from each database.

**a** **b**

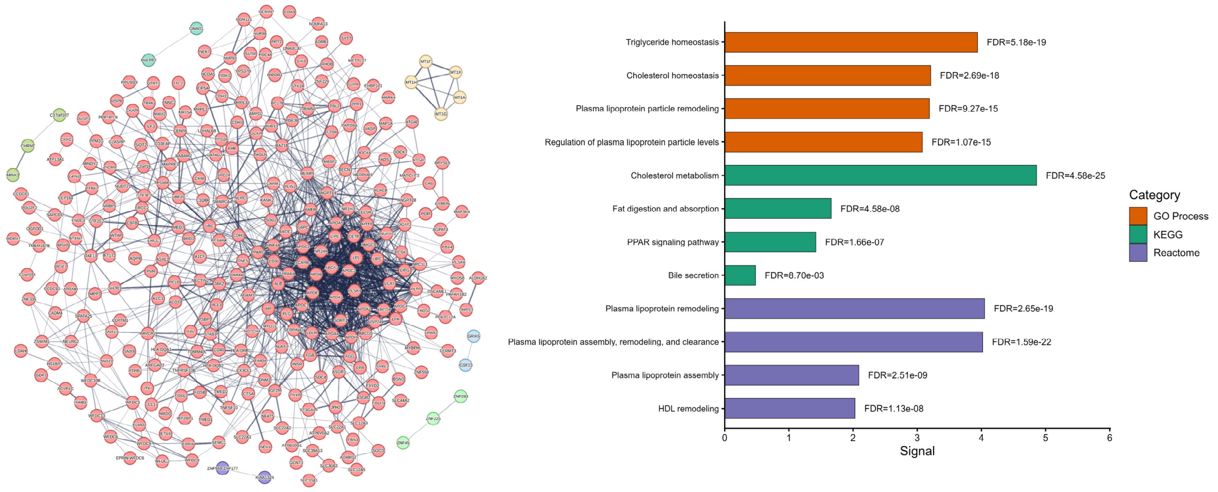

**Supplementary Figure 13 | Protein-protein interaction network and functional enrichment analyses of protein-coding genes identified from coding and noncoding analyses associated with NMR Lipoprotein subclasses (including composition) traits.** We applied Bonferroni-corrected genome-wide significance threshold of $\alpha = 0.05/(20{,}000 \times 7) = 3.57 \times 10^{-7}$ for protein-coding genes. Orphan genes (i.e., genes without connections in the PPI network) were hidden. **a** PPI network constructed from the identified protein-coding genes. **b** Significantly enriched pathways identified through Gene Ontology (GO), KEGG, and Reactome analyses, displaying the top four enriched pathways from each database.

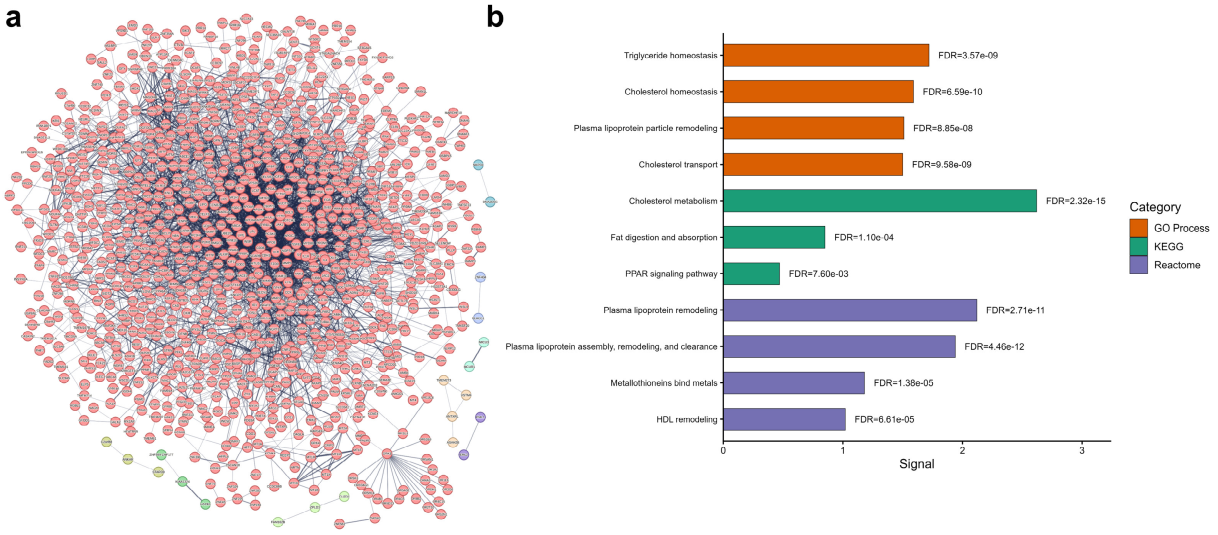

**Supplementary Figure 14 | Protein-protein interaction network and functional enrichment analyses of protein-coding genes identified from coding and noncoding analyses associated with NMR Amino acids traits.** We applied Bonferroni-corrected genome-wide significance threshold of $\alpha = 0.05/(20{,}000 \times 7) = 3.57 \times 10^{-7}$ for protein-coding genes. Orphan genes (i.e., genes without connections in the PPI network) were hidden. **a** PPI network constructed from the identified protein-coding genes. **b** Significantly enriched pathways identified through Gene Ontology (GO), KEGG, and Reactome analyses, displaying the top four enriched pathways from each database.

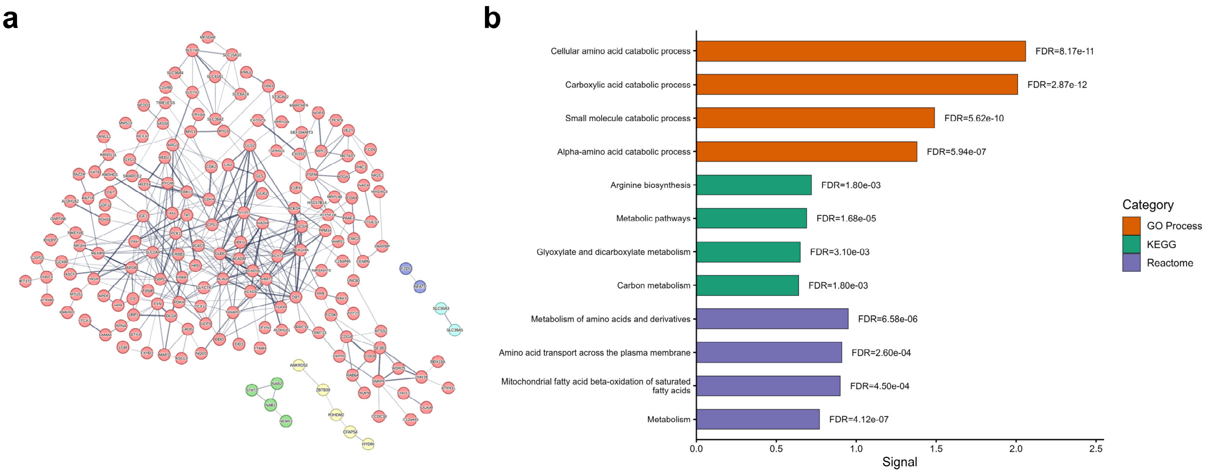

**Supplementary Figure 15 | Protein-protein interaction network and functional enrichment analyses of protein-coding genes identified from coding and noncoding analyses associated with NMR Others traits.** We applied Bonferroni-corrected genome-wide significance threshold of $\alpha = 0.05/(20{,}000 \times 7) = 3.57 \times 10^{-7}$ for protein-coding genes. Orphan genes (i.e., genes without connections in the PPI network) were hidden. **a** PPI network constructed from the identified protein-coding genes. **b** Significantly enriched pathways identified through Gene Ontology (GO), KEGG, and Reactome analyses, displaying the top four enriched pathways from each database.

**a** **b**

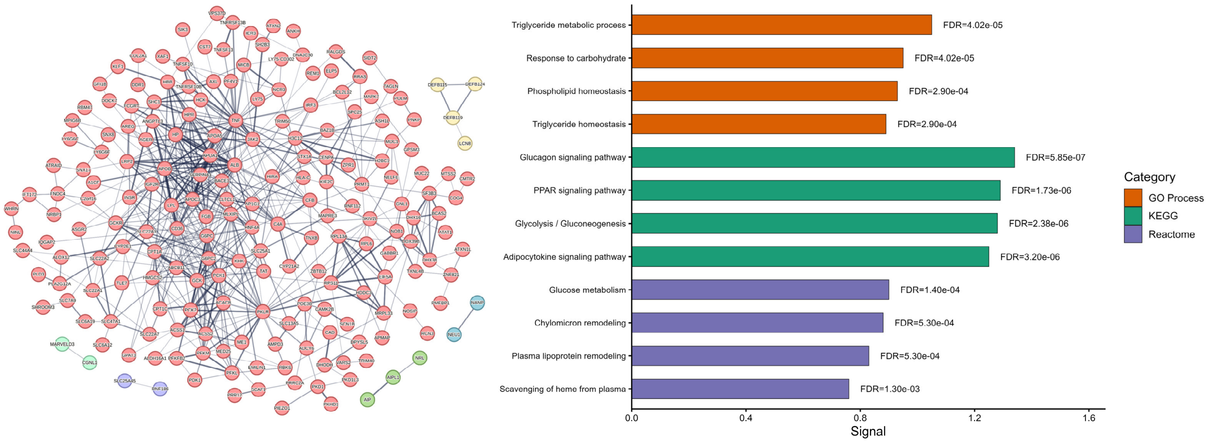

**Supplementary Figure 16 | Cloud cost (£) and computation time (in hours) of *STAARpipeline* (v0.9.8), *STAARpipeline* (v0.9.9), and *STAARpipelinePheWAS* (v0.9.7.1) for rare coding and noncoding variant set analyses using UK Biobank 500K WGS data.** Variant set analyses for rare variants (minor allele frequency < 1%) included gene-centric analysis of protein-coding genes across seven coding functional categories, seven noncoding functional categories, and rare variants in noncoding RNA (ncRNA) genes. Cloud costs are based on spot instances. Computation times for *STAARpipeline* v0.9.9 and *STAARpipelinePheWAS* v0.9.7.1 were estimated using mem3_ssd1_v2_x32 (30 cores), whereas *STAARpipeline* v0.9.8 used mem3_ssd1_v2_x48 or mem3_ssd1_v2_x64 instances with varying core counts. For *STAARpipelinePheWAS*, costs and computation time were benchmarked for simultaneous analyses of 5, 10, or 20 traits; per-trait costs or computation times are shown. Costs and computation times for variant set analyses, gene-centric coding, gene-centric noncoding, and ncRNA analyses are shown in red, blue, yellow, and green, respectively. (**a**) Costs for quantitative traits. (**b**) Costs for imbalanced case-control traits. (**c**) Computation times for quantitative traits. (**d**) Computation times for imbalanced case-control traits.

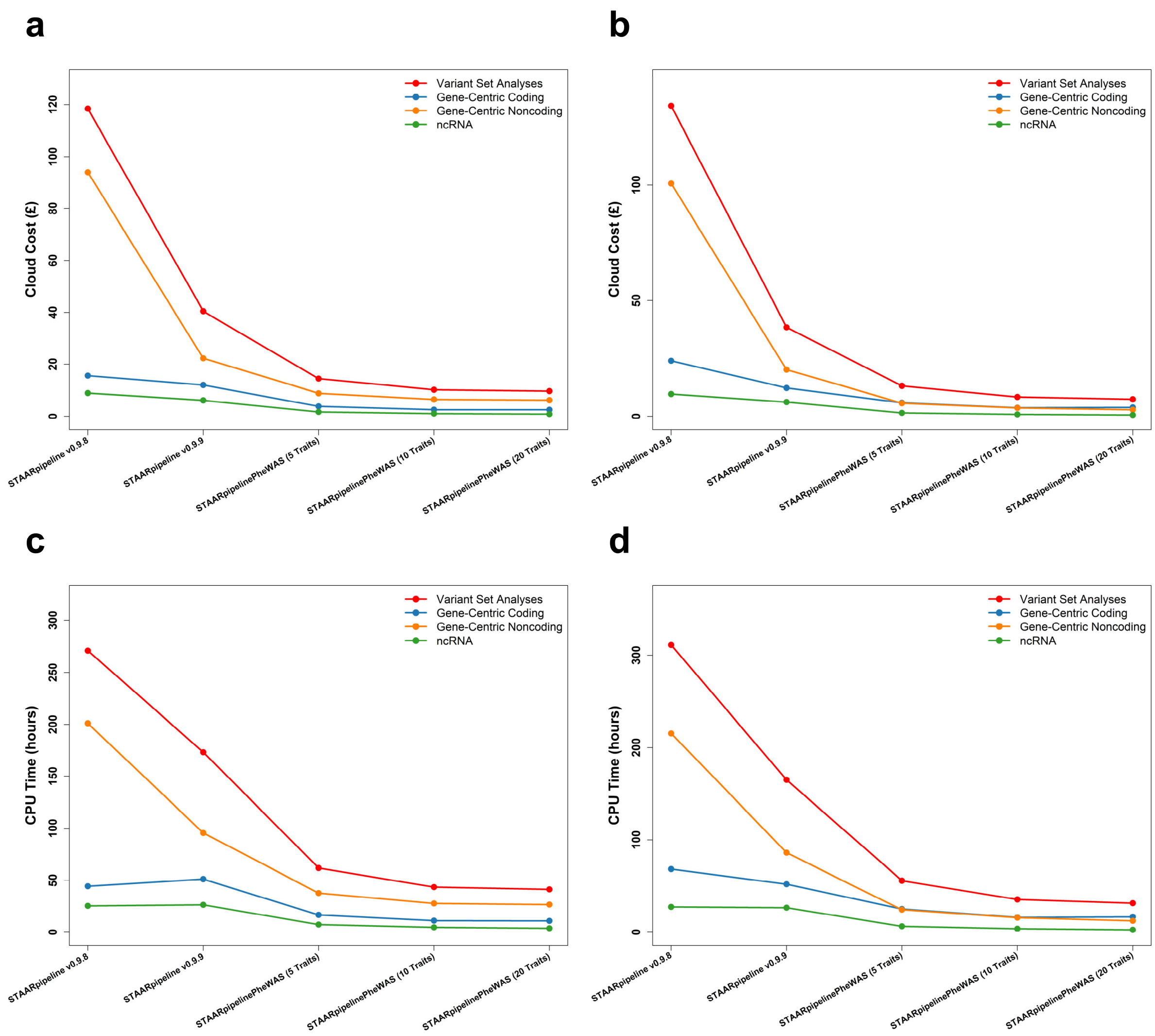
a
b
c
d
Variant Set Analyses
Gene-Centric Coding
Gene-Centric Noncoding
ncRNA
Cloud Cost (£)
CPU Time (hours)
STAARpipeline v0.9.8
STAARpipeline v0.9.9
STAARpipelinePheWAS (5 Traits)
STAARpipelinePheWAS (10 Traits)
STAARpipelinePheWAS (20 Traits)
